# Predicting the mutational drivers of future SARS-CoV-2 variants of concern

**DOI:** 10.1101/2021.06.21.21259286

**Authors:** M. Cyrus Maher, Istvan Bartha, Steven Weaver, Julia di Iulio, Elena Ferri, Leah Soriaga, Florian A. Lempp, Brian L. Hie, Bryan Bryson, Bonnie Berger, David L. Robertson, Gyorgy Snell, Davide Corti, Herbert W. Virgin, Sergei L. Kosakovsky Pond, Amalio Telenti

## Abstract

SARS-CoV-2 evolution threatens vaccine- and natural infection-derived immunity, and the efficacy of therapeutic antibodies. Herein we sought to predict Spike amino acid changes that could contribute to future variants of concern. We tested the importance of features comprising epidemiology, evolution, immunology, and neural network-based protein sequence modeling. This resulted in identification of the primary biological drivers of SARS-CoV-2 intra-pandemic evolution. We found evidence that resistance to population-level host immunity has increasingly shaped SARS-CoV-2 evolution over time. We identified with high accuracy mutations that will spread, at up to four months in advance, across different phases of the pandemic. Behavior of the model was consistent with a plausible causal structure wherein epidemiological variables integrate the effects of diverse and shifting drivers of viral fitness. We applied our model to forecast mutations that will spread in the future, and characterize how these mutations affect the binding of therapeutic antibodies. These findings demonstrate that it is possible to forecast the driver mutations that could appear in emerging SARS-CoV-2 variants of concern. This modeling approach may be applied to any pathogen with genomic surveillance data, and so may address other rapidly evolving pathogens such as influenza, and unknown future pandemic viruses.

## Introduction

SARS-CoV-2 evolution presents an ongoing challenge to public health. Many mutations have arisen in the SARS-CoV-2 genome as the pandemic has progressed. Most of these changes are expected to be neutral, with no functional or antigenic significance. Understanding the relative importance of mutations in viral proteins, particularly those of relevance for antiviral immunity, is therefore key to allocating preparedness efforts. Mutations in the viral Spike protein have received particular attention, because Spike is the target of antibody-mediated immunity and is the primary antigen in current vaccines^1^. As of April 24^th^, 2021, more than 6,200 distinct amino acid substitutions, insertions or deletions have been reported in Spike^2^. These mutations occur at all but two positions in the protein, in different combinations, creating over 45,000 unique Spike protein sequences. A small subset of these mutations are components of either “Variants of Interest” (VOIs) or “Variants of Concern” (VOCs), as classified by the Centers for Disease Control^3^. The distinction between VOIs and the higher alert VOCs is whether a negative clinical impact is suspected or confirmed^3^.

Early and objective identification of the key Spike amino acid changes contributing to future putative VOI/VOCs would be a boon to public health strategy. Such predictions could enhance the identification of liabilities for antibody-based therapeutics, vaccines and diagnostics. Predicting future mutations in variants that spread would extend the time available to develop proactive responses at earlier stages of spread. It would also complement existing forecasting efforts which seek to predict overall SARS-CoV-2 incidence, hospitalizations, and death over time^4–6^. As an indication of the need for mutation-centered models, the CDC aggregates results from 25 models that predict the number of new COVID-19 cases^7^. To our knowledge, no comparable models are in use for predicting SARS-CoV-2 mutations contributing to VOI/VOCs.

There is a robust and expanding set of data characterizing the features of amino acid mutations occurring in viral variants of SARS-CoV-2. Studies have identified the emergence of new variants with altered biological and/or antigenic properties^8–10^ and characterized them using low throughput methods^11,12^. Deep mutational scanning elucidates the *in vitro* biological effects of all single site amino acid substitutions^13–15^. Others have characterized the distribution of immunodominant sites across the viral proteome^16,17^ and estimated the fitness of viral sequences using neural natural language processing (NLP) applied to protein sequences^18^.

We seek here to build upon these data to forecast the mutations that will spread in the near future. We hypothesized that this would also allow us to simultaneously identify the dominant biological drivers of viral evolution over time. These two goals are mutually reinforcing: the features that are most useful for forecasting can be inferred as measuring viral fitness. Conversely, a better understanding of evolutionary dynamics can make modeling more accurate and robust. To accomplish these goals we (i) described patterns of rapid mutation spread both globally and within the United States, (ii) elucidated the relative predictive importance of amino acid mutational features comprising immunity, transmissibility, evolution, language model, and epidemiology; (iii) utilized data from previous waves to train and back-test a forecasting model that anticipates future spreading mutations, and (iv) illustrated how forecasted mutations could differentially affect clinical antibodies.

### A working definition for spreading amino acid mutations

For the purpose of developing the models, we defined “spreading” amino acid mutations as a specified fold change in frequency across multiple countries, comparing time windows before and after a chosen date (**Fig. 1**). We used over 900,000 SARS-CoV-2 Spike sequences available from GISAID^2^ as of April 24^th^, 2021 to characterize mutational spread within recognized and potential VOCs/VOIs globally and regionally within the United States. As a baseline analysis, we examined growth during the third wave of the pandemic (the three months after Nov. 1; **Fig. 1A**), using the three months prior as reference.

**Figure 1.**
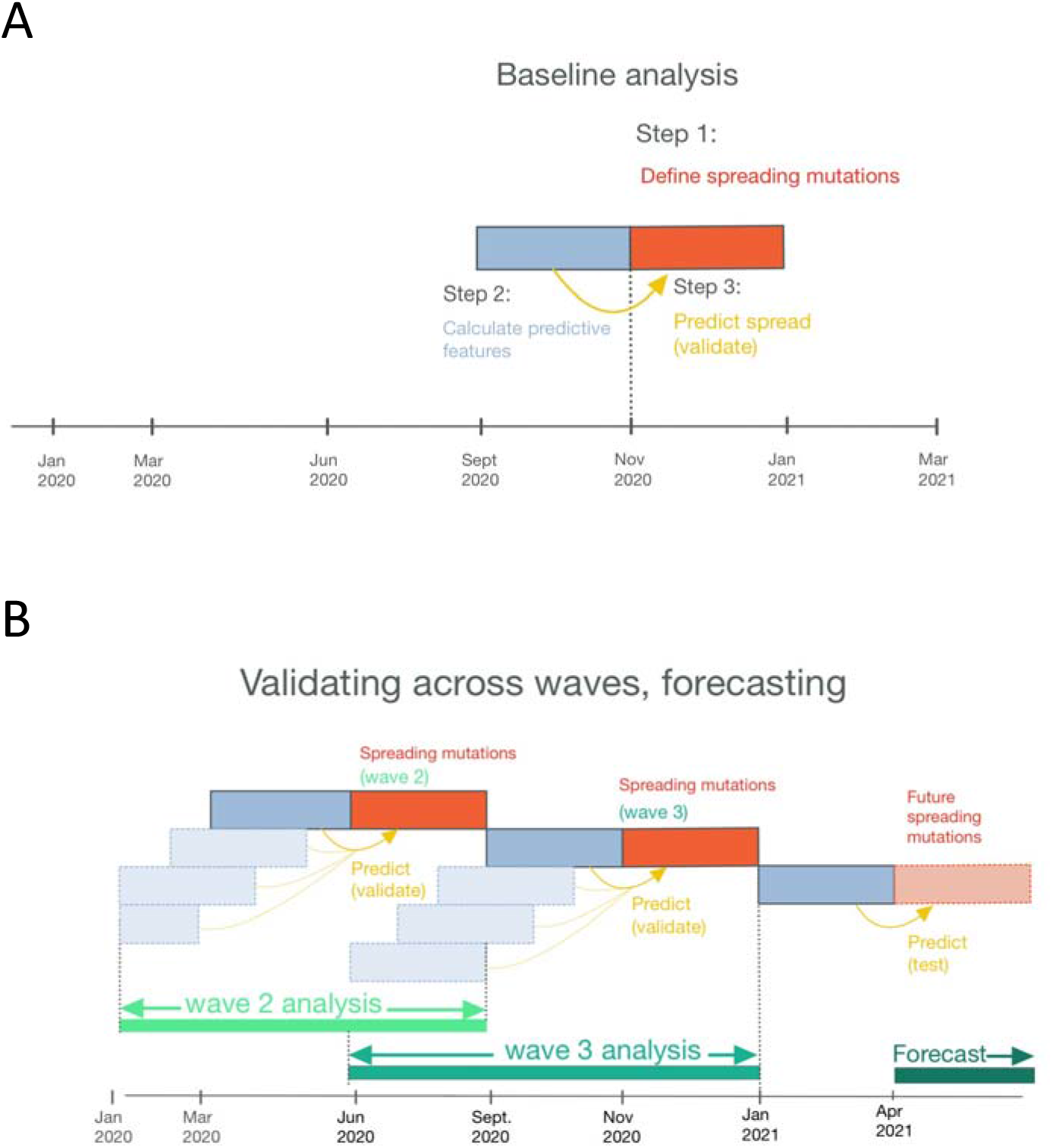
Baseline study design. **(A)** The core analysis consists of three steps. First, we create a working definition for spreading mutations. Second, we calculate features that can predict future spread using a window of prior data. Third, we construct models on training data, run prediction of future spread, and interpret the results. **(B)** For wave 2 and wave 3, we conduct the same steps as A: defining spreading mutations, calculating predictive features, and predicting spread. To assess the possible lead time provided by our model, we calculated predictive features at 1, 2, 3, and 4 months in advance. To query robustness, we looked at predictions for both wave 2 and wave 3. Finally, we generated predictive features based on contemporary data and used our model to forecast the mutations that could spread in the future.

We used Fisher’s exact test for frequency fold-change per country, adjusted for multiple comparisons, to identify a list of potentially spreading mutations. Within each country, we tabulated the number of sequences containing the mutation of interest, versus those that did not; in the three months before and after a date of interest (**Fig. 1A**). This was summarized as a 2×2 table per country, from which we calculated a fold change and an associated comparison-adjusted p-value. Mutations with a significant adjusted p-value from any country were retained. The number of comparisons for the adjustment was conservatively defined as the number of countries times the number of observed mutations worldwide. To account for violations in test assumptions (e.g., correlated counts due to biased sampling), and to enrich for the most concerning mutations, this set was further filtered using the following empirical criteria: (i) a fold change (FC) from baseline of at least 10.0 in at least one country, (ii) FC of at least 2.0 across three or more countries, (iii) a minimum global frequency of 0.1% in the later time window. We highlight that the sequences used to calculate fold change from baseline and minimum frequency were all collected after those used for model training or feature calculation, with no overlap or interleaving between the two datasets.

This definition of spreading mutations captured the current expansion of VOI/VOCs globally (**Fig. 2**) as well as the growth of a number of lesser-known mutations (**Suppl. Fig. S1A**). For example, R346K and V367F mutations, increase Spike expression and contribute to immune escape^14,19^. R346K increased 7x in Switzerland, 8x in Austria, and 21x in Chile. V367F frequency increased at least 8x in the United Kingdom and Switzerland, 12x in Spain, and 36x in Canada. The broadest geographic increases were observed for P681R, which increased over 4x in 15 countries, and over 20x in 7 countries. P681R adds a basic amino acid adjacent to the Spike furin cleavage site and enhances the fusion activity of Spike^20^.This mutation is now dominant in the B.1.617.1/.2/.3 lineages. Thus, we detected increases in the frequency of this mutation well before the current wave of VOC-associated disease in India. We detected additional spreading mutations (N501T, D138H, and W152L) at sites in the Spike already associated with another spreading set of mutations (N501Y, D138Y and W152C). We confirmed that N501T, like N501Y, also improves ACE2 binding of Spike^21^.

**Figure 2.**
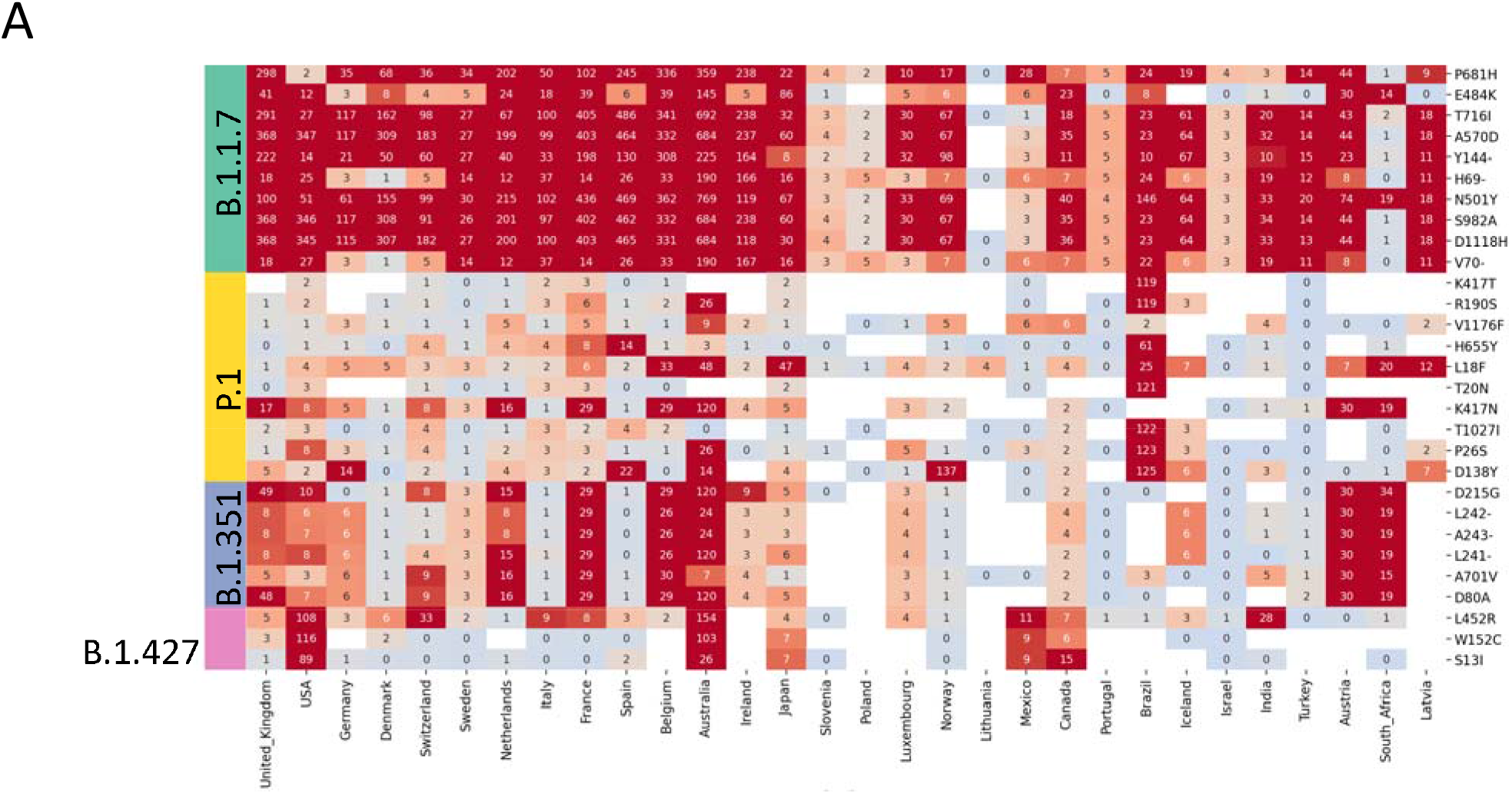
Describing spreading mutations in the third wave. The working definition of spreading mutations captures the expansion of variants of concern during wave 3 **(A)** at the country level and **(B)** at the state level within the United States. Leading variants of concern are denoted by colored bars on the left-hand side. Green: Alpha B.1.1.7; Yellow: Gamma P.1; Blue: Beta B.1.351; Pink: Epsilon B.1.427/B.4.429. The emergence of E484K in association with Alpha B.1.1.7 is depicted in panel B. The Epsilon B.1.427/B.4.429 is observed across multiple US states. Previously unidentified potential spreading mutations are denoted by a beige colored bar on the left-hand side. The x-axis (countries or states) are ordered left to right according to decreasing number of GISAID submissions being represented.

Together, these data indicate that our definition of globally spreading mutations in wave 3 of the pandemic (**Fig. 1B**) identifies pandemic-relevant changes in Spike protein function and/or antigenicity. We next analyzed regional patterns of mutation spread within the United States (**Fig. 2B**), such as the spread of mutations carried by both Alpha (B.1.1.7) and Epsilon (B.1.427/ B.1.429) VOCs. We found that most of the increase in VOI/VOC mutations was observed in 14/50 states. Michigan, Florida, and Texas showed the most pronounced fold changes in mutations. We identified a set of less well-known mutations that appeared to be spreading in some regions (**Fig. 2B**). For example, T478K expanded over 60x in Texas (and 41x in neighboring Mexico). This mutation also increased at least 10x in Washington, California, and Oregon. Of note, T478K increases *in vitro* Spike expression and ACE2 binding^14^. We also analyzed regional patterns of mutation spread within the United States (**Suppl. Fig. S1A**), such as the spread of mutations carried by both Alpha (B.1.1.7) and Epsilon (B.1.427/B.1.429) VOC. We conclude that the proposed statistical criteria successfully identified the dynamics of mutations in SARS-CoV-2 variants, and detected the spread of less obvious mutations globally and within the United States.

### Biological and epidemiological features of mutations that spread

We next determined which features of amino acid mutations are useful for predicting the emergence in successful viral variants. Mutations were first ranked directly using each feature with no model fitting step (see **Supplemental Methods**). Since the Receptor Binding Domain (RBD) region of Spike had the most complete data, our initial analysis focused on RBD. Within the RBD, we found that ACE2 binding affinity was a slightly better predictor of mutation spread (area under the receiver operator characteristic curve, AUROC=0.84; **Fig. 3A**) than changes in in vitro expression of Spike mutants (AUROC=0.81; **Suppl. Fig. S2A**). Among measures of immune escape, the binding contributions of known antibody epitopes (antibody binding score; see materials and methods) to anti-SARS-CoV-2 antibodies were predictive of mutation spread (AUROC=0.75; **Fig. 3A**). We found that Natural Language Processing (NLP) scores for sequence plausibility (grammaticality)^18^ were similarly predictive (AUROC=0.77; **Fig. 3A**)^18^. We did not find that CD4^+^ or CD8^+^ T-cell immunogenicity offered significant explanatory power for mutation spread (AUROC=0.59; Suppl. **Fig. S2A**). The best evolutionary feature for prediction of spread (AUROC=0.91; **Fig. 3A**) was obtained from Fixed Effects Likelihood (FEL^23^) from the Hyphy package [http://www.hyphy.org]^24^ which tests for pervasive negative or positive selection across the internal branches of a phylogenetic tree. Positive selection occurs when new mutations contribute to higher fitness than wild-type, leading to a relative increase in amino acid diversity over time, whereas negative selection reduces the frequency of non-wild-type amino acids at that site.

**Figure 3.**
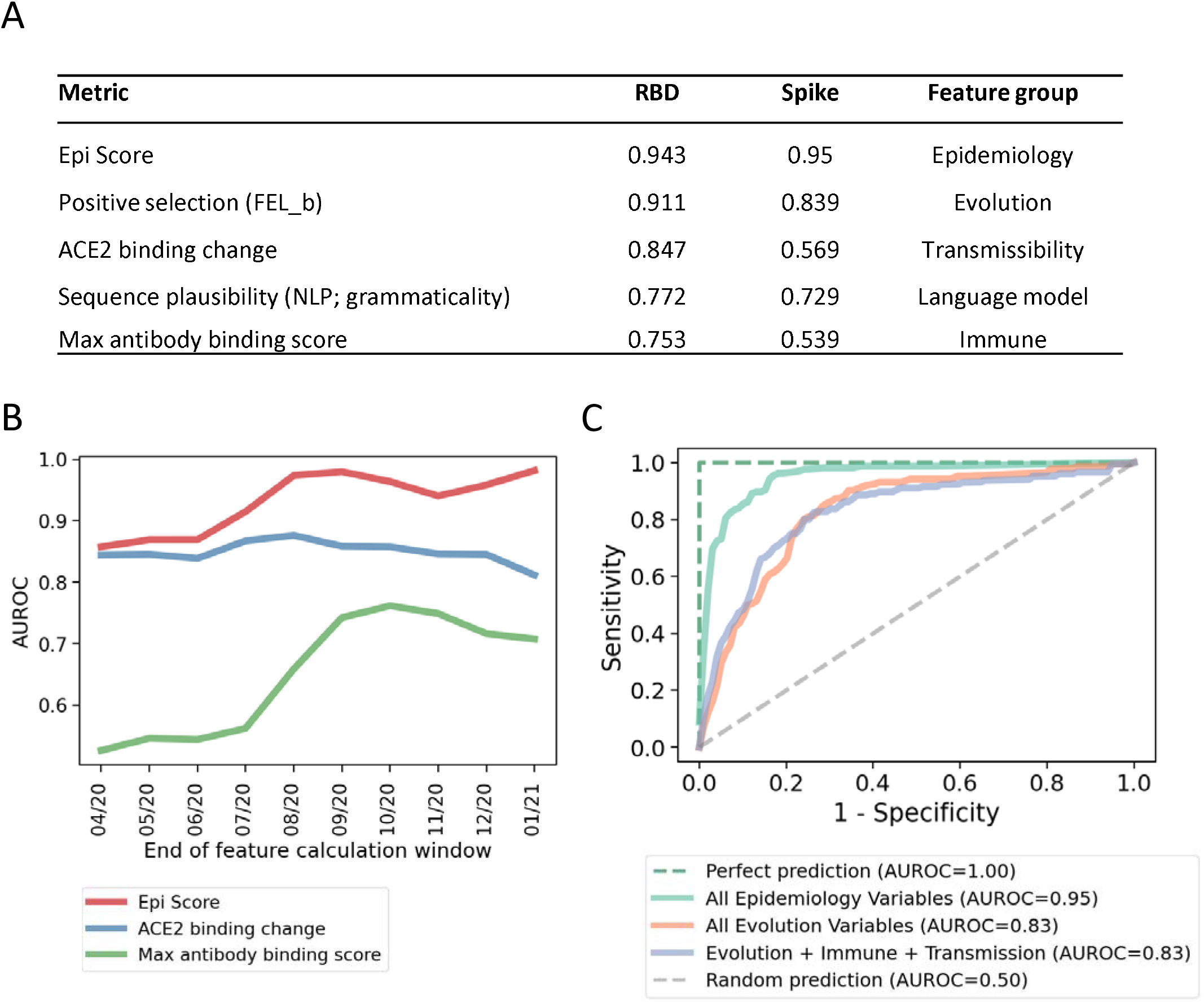
Predictors of mutation spread. Each variable was used to directly rank mutations for future spread. **(A)** The most predictive variables within each feature group (see Table 1 and Suppl. Table S1) are ranked by performance within the receptor binding domain (RBD), where the most data are available. Scores are Area Under the Receiver Operating Characteristic curve (AUROC). **(B)** RBD classification accuracy over time for the top GISAID-based feature (Epi score), and the top transmission and immune variables (Table 1). AUROCs in panel B are smoothed with a rolling window of two analysis periods. **(C)** Performance for identifying which mutations will spread during the baseline analysis period (see Fig 1A).

The highest predictive performance, however, was obtained from epidemiological features, i.e., variables which more directly take into account sampled mutation counts (**Table 1**). The most predictive variable in this feature category was “Epi Score”, the exponentially weighted mean ranking across the other epidemiological variables (AUROC=0.94). We interpret this metric to capture both lineage expansion and recurrent mutation that occurs in multiple variant lineages by convergent evolution. Both of these aspects of spread suggest that a specific mutation is contributing to virus fitness. We note that the utility of recurrent mutation signals is consistent with recent findings that convergent evolution plays a significant role in SARS-CoV-2 adaptation^25^. Outside of the RBD, there is less experimental annotation of amino acid mutations. As observed for the RBD alone, within Spike we obtained the best predictive performance with evolutionary (AUROC=0.84) and epidemiologic (AUROC=0.94) measures (**Fig. 3A**). The performance of other feature sets is presented in **Suppl. Fig. S3**. Outside of the RBD, there is less experimental annotation of amino acid mutations. CD4^+^ and CD8^+^ T-cell immunogenicity had little explanatory power across the full-length Spike sequence (max AUROC of 0.54). Language model grammaticality performance was slightly reduced in predictive impact compared to the same measure from RBD alone, with an AUROC of 0.73. As observed for the RBD alone, within Spike we obtained the best predictive performance with evolutionary (AUROC=0.84) and epidemiologic (AUROC=0.94) measures (**Fig. 3A**). The performance of other feature sets is presented in **Suppl. Fig. S3**.

**Table 1.** Summary of analytical features. A total of 48 parameters for 14 variables were created for 5 feature groups. These features capture evolutionary, immune, epidemiologic, transmissibility, and language model predictors of mutation spread. A detailed description of all parameters is included in Suppl. File 1.

| Feature group | Variable | Meaning | Source or reference | Number of parameters |
| --- | --- | --- | --- | --- |
| Evolution | Positive selection (FEL, MEME) | Parameters from Fixed Effects Likelihood (FEL) and Mixed Effects Model of Evolution (MEME) | HyPhy<sup>24</sup> | 11 |
|  | Codon-SHAPE | RNA SHAPE constraint | Manfredonia et al. 2020<sup>44</sup> | 3 |
|  | Viral entropy | Shannon entropy at each codon position for an amino acid site | This work | 3 |
| Immune | CD8 epitope escape | The frequency of SARS CoV-2 mutations in cytotoxic lymphocyte (CTL) epitopes | Agerer et al. 2021<sup>16</sup> | 1 |
|  | CD8 response | The percent and average CD8+ T-cell response to an epitope in patients | Tarke et al. 2021<sup>42</sup> | 2 |
|  | CD4 response | The percent and average CD4+ T-cell response to an epitope in patients | Tarke et al. 2021<sup>42</sup> | 2 |
|  | Antibody binding score | The estimated percent contribution of a site to binding of the indicated antibody, as estimated by Molecular Operating Environment (MOE) | This work | 17 |
|  | Maximum escape fraction in vitro | The maximum escape fraction across all conditions for that mutation | Greaney et al. 2021<sup>21</sup> | 1 |
| Epidemiology | Variant frequency | The percent of sequences with the mutation | Calculated from GISAID<sup>2</sup> | 1 |
|  | Fraction of unique haplotypes | The fraction of unique Spike haplotypes in which a mutation is observed | Calculated from GISAID<sup>2</sup> | 1 |
|  | Number of countries | The number of countries where it has been observed. | Calculated from GISAID<sup>2</sup> | 1 |
|  | Epi Score | The exponentially weighted mean rank across the other epidemiology variables | Calculated from GISAID<sup>2</sup> | 1 |
| Transmissibility | RBD expression change | Change in RBD expression due to the mutation | Starr et al. 2020<sup>14</sup> | 1 |
|  | ACE2 binding change | The change in binding affinity for ACE2 | Starr et al. 2020<sup>14</sup> | 1 |
| Language model | Language model | Grammaticality and semantic change of a mutation | Hie et al. 2021<sup>18</sup> | 2 |

We next sought to interrogate the robustness of this approach to changes in the underlying drivers of SARS-CoV-2 evolution. For example, it has been hypothesized that selection due to immune pressure has increased with time as more individuals became immune through infection or vaccination^25^. For example, the Gamma P.1 lineage is thought to have spread rapidly in Brazil largely due to immune selection in a population with high seroprevalence^26^. We measured the predictive performance of antibody binding scores, which quantify the predicted percent contribution of each Spike site to antibody affinity. We take this metric as a proxy for B-cell immunodominance (**Table 1**)^27^. Taking the maximum of this value across antibodies at a given site yields the maximum antibody binding score. The predictiveness of this metric increased from nearly uninformative early in the pandemic (p-value for difference from random=0.53), to an AUROC of 0.75 (p<1e-4; **Suppl. Fig. S2C**) for predicting spreading mutations during the third wave of the pandemic (**Fig. 3B**).

A similar analysis for all variables in both Spike and RBD is presented in **Suppl. Fig. S2A** and **S2B**. Throughout this transition from lower to higher levels of immune selection, we found that epidemiological features maintain their performance, achieving an AUROC of 0.98 for the final evaluation period (**Fig. 3B**). A summary of performance for all features across time and within the RBD and across full-length Spike can be found in **Suppl. Fig. S2A** and **S2B**.^24^

We next trained a model to predict spreading mutations using sets of features identified above. We employed logistic regression with baseline features as inputs. Within each feature set, the features used for prediction were selected using forward feature selection, cross-validated within each training dataset. To minimize correlation between training and test amino acid mutations through shared haplotype structure, model training was arranged so that mutations from the same phylogenetic clade were never split across the training and test datasets, thus minimizing information leakage. For the third wave of infections at one month of anticipation (**Fig. 1A**), the best predictors were positive selection features (AUROC=0.83) and epidemiologic features (AUROC=0.95; **Fig. 3C**). Immunity and transmissibility features did not improve predictive power of positive selection features (AUROC=0.83). We did not find additional variables that improved upon the performance of epidemiological features. The performance of the trained model was comparable to the performance of the Epi Score (**Fig. 3A** vs. **Fig. 3C**). Therefore, to simplify reproducibility and further minimize the risk of overfitting, we used the Epi Score to predict mutation spread going forward.

In summary, immunity, transmissibility, evolution, language model, and epidemiologic features all effectively predict mutation spread and that the methodology accommodates to and captures changes to the underlying selective forces over the course of the pandemic. We find that epidemiologic features in particular display superior accuracy and robustness over time.

### Examining global dynamics and the emergence of VOCs

To determine whether local or global dynamics drive mutation spread, we examined whether spreading mutations in the United States were better predicted by global or US-only epidemiological values. For this, we first tested the performance of the Epi Score to forecast spreading mutations in both the second and third wave; with one, two, three, and four months of anticipation (**Fig. 1B)**. We found that mutations were predicted with an AUROC above 0.85 at least two months in advance, in both wave 2 and wave 3 both within the United States and globally. Prediction of mutations in Wave 2 was still better than random four months in advance, despite only having access to the fewer than 600 viral sequences that were available in January and February of 2020. In Wave 3, we observed AUROCs of 0.87 for predicting both United States and globally spreading mutations four months in advance (**Fig. 4A)**. We found that global epidemiology metrics were best overall and were generally more predictive of state-level mutation spread than the state-level metrics themselves (**Suppl. Fig. S4**).

**Figure 4.**
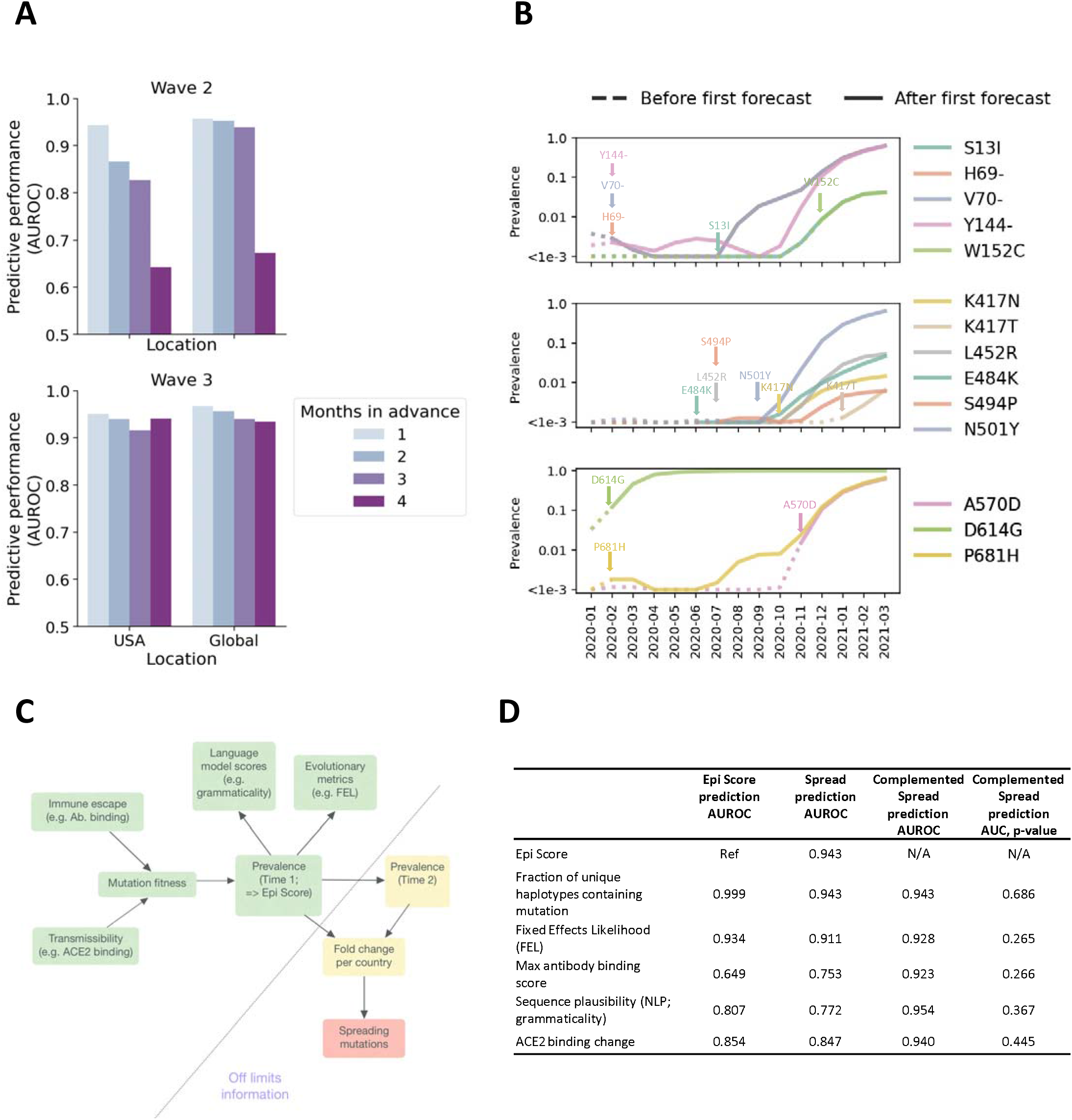
Early detection and causal mediation of spreading mutations. **(A)** Performance (AUROC) in predicting spreading mutations in the second and third waves of the epidemic (see Fig. 1). Predictions are generated using a three-month window of data, preceding the wave by 1-4 months. **(B)** Depiction of where in their growth trajectories VOC mutations were first forecast to spread. Dotted lines denote the part of the curve where the variant had not yet been forecast to spread. Solid lines denote the period after first forecast. A version of this plot with mutations grouped by lineage can be found in Suppl. Fig S4. To reduce overplotting, mutations are plotted in genomic order, but split into panels by genomic region: NTD, RBD, and other regions. **(C)** Model: viral fitness (determined by, e.g. changes to transmissibility and immune escape) drives viral prevalence at time 1 (as measured by global frequency, and geographic and haplotype distribution), which is captured by Epi score. Language model score or evolutionary metrics are summaries of GISAID data and therefore are shaped by mutation prevalence. Prevalence at time 1 predicts prevalence at time 2, which ultimately leads to mutation being defined as spreading. Therefore, prevalence at time 1 (as captured by Epi Score) mediate the effects of the biological variables that enhance viral fitness through transmissibility or escape adaptation. **(D)** To quantitatively test for mediation, we assessed whether variables were better at predicting top mutations that are in the top 200 Epi Score, compared to spreading mutations for time 2 versus time 1. Within the RBD, the major variables from each group generally predict Epi Score better than they predict spreading mutations. One exception was antibody binding score, which is discussed in the main text. In support of mediation, the analysis found little complementarity, as measured by the AUROC, when combining each variable with the most effective epidemiological variable.

To illustrate the practical utility of Epi Score using global features, we performed the sliding window analysis to assess how early we would have been able to forecast the spread of Spike mutations contained in current CDC VOCs. To be conservative, we counted the date that a mutation was first forecast as the earliest date at which it was predicted to spread in two subsequent analysis periods. For example, consider a mutation that was predicted to spread in June, October, and November. Since the mutation was not predicted to spread in July, the June date would be disregarded and the earliest forecasted date would instead be October. The chosen cutoff of the 200 top scoring mutations corresponded to a historical specificity between 93% and 97% across the sliding window period, with a specificity of 97% in the most recent period (**Suppl. Fig. S5**).

We next retrospectively examined when we would have been able to predict the spread of current VOC Spike amino acid mutations (**Fig. 4B** and **Suppl. Fig. S6**). Since D614G was already highly prevalent before sufficient data were available to inform predictions, early forecasting was not meaningful for this mutation. The rise of A570D was also rapid enough that it was not forecast until it reached 1.3% frequency. Even including these mutations, we could predict VOC mutations an average of more than 5 months in advance of them reaching 1% global frequency. For the mutations forecasted prior to reaching this threshold, the average frequency at the time of first forecast was 0.16% (**Suppl. Table S1**). We repeated this analysis for the more recently emerged Beta lineages B.1.617.1, .2 and .3 first discovered in India. We find that the L452R mutation from this lineage was first forecast in July of 2020, while the P618R was first forecast in October of 2020. The E484Q mutation associated with B.1.617.1 and .3 (presumably lost from B.1.617.2) was not forecast until March of 2021 (**Suppl. Table S2**). We conclude that the approach was robust enough to predict key mutations in the second and third waves of the pandemic several months in advance. Early warning of mutations in current VOCs and VOIs would have been possible before reaching worrisome levels of global spread.

### Understanding performance through a causal lens

Seeking to understand the high predictive performance of epidemiologic features, we constructed a directed acyclic graph to visualize the hypothesized causal relationships, and to probe whether relative trends in performance were consistent with the expectations that follow from this model (**Fig. 4C**). We proposed that epidemiologic features mediate the relationship between viral fitness and sets of mutations spreading. Our rationale was that if a mutation’s contribution to viral fitness was sufficient to drive it to appreciable prevalence at one time point (as measured by global frequency and geographic and haplotype distribution), it would likely drive it to higher prevalence in the future as well (unless it were outcompeted by a more fit adaptation, or the fitness landscape changes). This type of mediated relationship (fitness => current prevalence => future prevalence) implies that epidemiological prevalence features will capture information from both known and unknown drivers of selection.

While such an approach could also identify neutral hitchhiker mutations, averaging across all genetic backgrounds mitigates this risk. The utility of epidemiologic features for prediction extends beyond measuring fitness. Even for two adaptive mutations that confer the same fitness advantage, the one that is currently more prevalent has a higher likelihood of spreading further, since higher prevalence increases the influence of selection relative to genetic drift^28^. This line of reasoning predicts that epidemiologic variables that capture relative initial prevalence, will provide a robust measure of a mutation ultimate success. Epistasis, public policy, and a range of other factors could obscure this relationship. For example, otherwise adaptive mutation could be unsuccessful on an incompatible genetic background. Neutral mutations could also increase in frequency in areas due to an insufficient public health response. However, by focusing on amino acid mutations across viral variants and geographic locations, we were able to average these effects out to some extent.

If the presented causal model were reasonable, we would expect first that variables whose causal effects are mediated, as defined above, should predict epidemiologic variables at a comparable or even greater accuracy compared to spreading mutations. This is illustrated by comparing the first and second columns of **Fig. 4D**. We observed that, with the exception of the maximal antibody binding score, all top variables predict Epi Score better than they predict mutation spread. The lower predictiveness of maximal antibody binding score for Epi Score would be consistent with a slight time lag effect due to shifting evolutionary pressures.

A second criterion for mediation is that information from these variables should not significantly complement the predictiveness of the epidemiologic variables alone. We can assess this by comparing the AUROCs of two-variable models in column 3 of **Fig. 4D** with the AUROC for Epi Score alone (0.94). The only nominal AUROC increase for a complemented model was observed for NLP sequence plausibility (0.95). This improvement was not statistically significant (p=0.367). Similarly, we do not find statistically significant complementarity with Epi Score for any other variable, either within the RBD or across full length Spike (see supplemental section “Mediation Analysis”, **Suppl. Table S3)**.

Our examination of mediated causal relationships begins by assuming a causal graph based on prior knowledge. Such an approach is common to many causal inference methods^29^ and represents a well-understood limitation of these methods^29^. Therefore, we consider this as a tool to more systematically analyze the plausibility of our results. While it is generally difficult to verify the structure of proposed causal graphs, our findings support the concept that epidemiological variables mediate the effects of other classes of explanatory variables, and this may explain their high predictive accuracy.^26^

### Forecasting spreading mutations

Building upon the accurate prediction of spreading mutations in the second and third wave, the stability of performance in the face of changing selective dynamics, and the explainability of high predictive performance of epidemiologic features, we next leveraged Epi Score on the current data to forecast mutations that may contribute to VOIs and VOCs over the coming months. Since global metrics outperformed metrics restricted to the United States, even for forecasting within the United States, we focused on global forecasting. We considered shortening our feature calculation window to further mitigate the effects of shifting evolutionary dynamics. However, we found that longer feature calculation windows robustly improved performance across all prediction windows (**Suppl. Fig. S7**).

We present in **Table 2** a subset of 22 predicted mutations that do not belong to the canonical Alpha B.1.1.7, Beta B.1.351, Gamma P.1, or Epsilon B.1.427/B.1.429 VOC haplotypes, and obtained an Epi Score of at least 9.8 out of 10. A visualization of how the frequency of all forecast mutation have changed over time can be found in **Suppl. Fig. S8**. We find that most forecast amino acid mutations have demonstrated consistent increases in global frequency. A complete list of forecast mutations is presented in **Suppl. Table S4**. We posit that top forecast mutations should we considered for in depth analysis of emergent biological properties such as immune evasion (see below) and or greater transmissibility.

**Table 2.** Selected forecasted mutations. Included are forecasted mutations that are not associated to CDC variants of concern and score above 9.7 in Epi score. RBM = Receptor binding motif; NTD = N-terminal domain.

| Mutation | Epi Score | Spike region | Date first reported | Most prevalent lineage | Number of lineages | Counts (n) on VOCs |
| --- | --- | --- | --- | --- | --- | --- |
| S12F | 9.8 | Signal peptide + NTD | 2020-03 | B.1.1.7 | 122 | B.1.1.7 (761), B.1.351 (9),<br>B.1.427 (3), P.1 (70) |
| Q52R | 9.8 | Signal peptide + NTD | 2020-05 | B.1.525 | 27 | B.1.1.7 (50), B.1.427 (1) |
| S98F | 9.9 | Signal peptide + NTD | 2020-01 | B.1.221 | 166 | B.1.1.7 (4875), B.1.351 (57),<br>B.1.427 (14) |
| D138H | 9.8 | Signal peptide + NTD | 2020-02 | B.1.1.7 | 60 | B.1.1.7 (2765), B.1.427 (1) |
| L141- | 9.8 | Signal peptide + NTD | 2020-02 | A.2.5 | 153 | B.1.1.7 (50), B.1.351 (16),<br>B.1.427 (2), P.1 (16) |
| G142- | 9.8 | Signal peptide + NTD | 2020-02 | B.1.1.7 | 158 | B.1.1.7 (1070), B.1.351 (16),<br>B.1.427 (2), P.1 (17) |
| M153T | 9.8 | Signal peptide + NTD | 2020-01 | B.1.1.284 | 77 | B.1.1.7 (307), B.1.427 (1) |
| L189F | 9.8 | Signal peptide + NTD | 2020-05 | B.1.258.17 | 36 | B.1.1.7 (20), P.1 (1) |
| A222V | 10 | Signal peptide + NTD | 2020-02 | B.1.177 | 284 | B.1.1.7 (449), B.1.351 (22),<br>B.1.427 (45), P.1 (2) |
| A262S | 9.8 | Signal peptide + NTD | 2020-02 | B.1.177 | 98 | B.1.1.7 (91), B.1.351 (4), B.1.427<br>(1), P.1 (11) |
| S477N | 9.9 | RBM | 2020-01 | B.1.160 | 145 | B.1.1.7 (22), B.1.427 (1), P.1 (1) |
| T478K | 9.9 | RBM | 2020-04 | B.1.1.519 | 38 | B.1.1.7 (3), B.1.351 (1), B.1.427<br>(1) |
| S494P | 9.8 | RBM | 2020-03 | B.1.575 | 112 | B.1.1.7 (1184), B.1.427 (27) |
| Q675H | 9.9 | Unannotated 2 | 2020-02 | B.1.1.214 | 178 | B.1.1.7 (486), B.1.351 (40),<br>B.1.427 (2), P.1 (4) |
| Q677H | 9.9 | Unannotated 2 | 2020-01 | B.1.2 | 240 | B.1.1.7 (2048), B.1.351 (26),<br>B.1.427 (51), P.1 (8) |
| T732A | 9.8 | Unannotated 2 | 2020-04 | B.1.1.519 | 31 | B.1.1.7 (4), B.1.351 (1) |
| G769V | 9.8 | Unannotated 2 | 2020-03 | R.1 | 154 | B.1.1.7 (53), B.1.351 (16) |
| A845S | 9.8 | Fusion Peptides | 2020-03 | B.1.1.317 | 132 | B.1.1.7 (306), B.1.351 (102),<br>B.1.427 (1), P.1 (24) |
| F888L | 9.8 | Unannotated 3 | 2020-03 | B.1.525 | 20 | B.1.1.7 (5) |
| S939F | 9.8 | Heptad repeat 1 | 2020-02 | B.1.1.7 | 174 | B.1.1.7 (1140), B.1.351 (12),<br>B.1.427 (1), P.1 (2) |
| K1191N | 9.8 | Heptad repeat 2 | 2020-03 | B.1.1.7 | 94 | B.1.1.7 (8097), B.1.351 (3),<br>B.1.427 (59), P.1 (2) |
| V1228L | 9.8 | Unannotated 5 | 2020-01 | B.1.596 | 133 | B.1.1.7 (443), B.1.351 (2),<br>B.1.427 (9), P.1 (6) |

Finally, as an application of the forecasting analysis, we examined how forecasted mutations intersected with the binding sites of clinical antibodies. We found a wide variation in the number of forecasted mutations per antibody epitope (**Table 3**), ranging from 8 mutations for Celltrion’s CT-P59, to zero mutations for Vir-7831 (sotrovimab), which was designed to be robust to viral evolution by targeting a region that is conserved across coronaviruses^30^. As proof of concept, we focused our attention on the forecast S494P, a variant reported to have enhanced binding affinity to ACE2^31^, reduced neutralization by 3-5-fold in some convalescent sera^31^ and increasing association with the B.1.1.7 lineage. As predicted, we show in **Fig. 5** that S494P mutation decreases neutralization potential of clinical therapeutic antibodies: Ly-CoV555 (bamlanivimab), CT-P59 and to a lesser extent to REGN10933 (casirivimab). Outside of the RBD, we note that a significant proportion of the forecasted mutations (39%) occur in the signal peptide and N-terminal domain (NTD), despite comprising 23% of the Spike sequence. Mutations in the signal peptide can affect processing of the spike and result in altered structures in the NTD^1,32^. More generally, the NTD, is the focus of much investigation^33^, as it is subject of considerable humoral immunity and thus is a hot-spot for viral adaptation^1^.

**Table 3.** Forecasted mutations for therapeutic antibodies. Unfiltered forecasted mutations (including VOC mutations) were intersected with the binding epitopes of therapeutic monoclonal antibodies. Mutations were included if they were in sites contributing at least 1% of the total binding energy for a given antibody, as estimated by Molecular Operating Environment (MOE) program. Mutations present at less than 1% global frequency in the most recent three months are presented in bold. Mutations already present at more than 1% global frequency in the most recent three months are presented in italics.

| Clinical therapeutic antibody | Forecasted mutations in epitopes |
| --- | --- |
| Vir-7831 (Sotrovimab) | None |
| LY-CoV016 (etesevimab) | *K417N*, **K417T** |
| REGN10987 (Imdevimab) | *N439K*, **N440K**, **G446V** |
| BD-368-2 | *L452R*, **L452Q**, *E484K*, **E484Q**, **F490S** |
| LY-CoV555 (Bamlanivimab) | *L452R*, **L452Q**, *E484K*, **E484Q**, **F490S**, **S494P** |
| REGN10933 (Casirivimab) | *K417N*, **K417T**, *S477N*, *T478K*, *E484K*, **E484Q**, **F490S** |
| CT-P59 | *K417N*, **K417T**, *L452R*, **L452Q**, *E484K*, **E484Q**, **F490S**, **S494P** |

**Figure 5.**
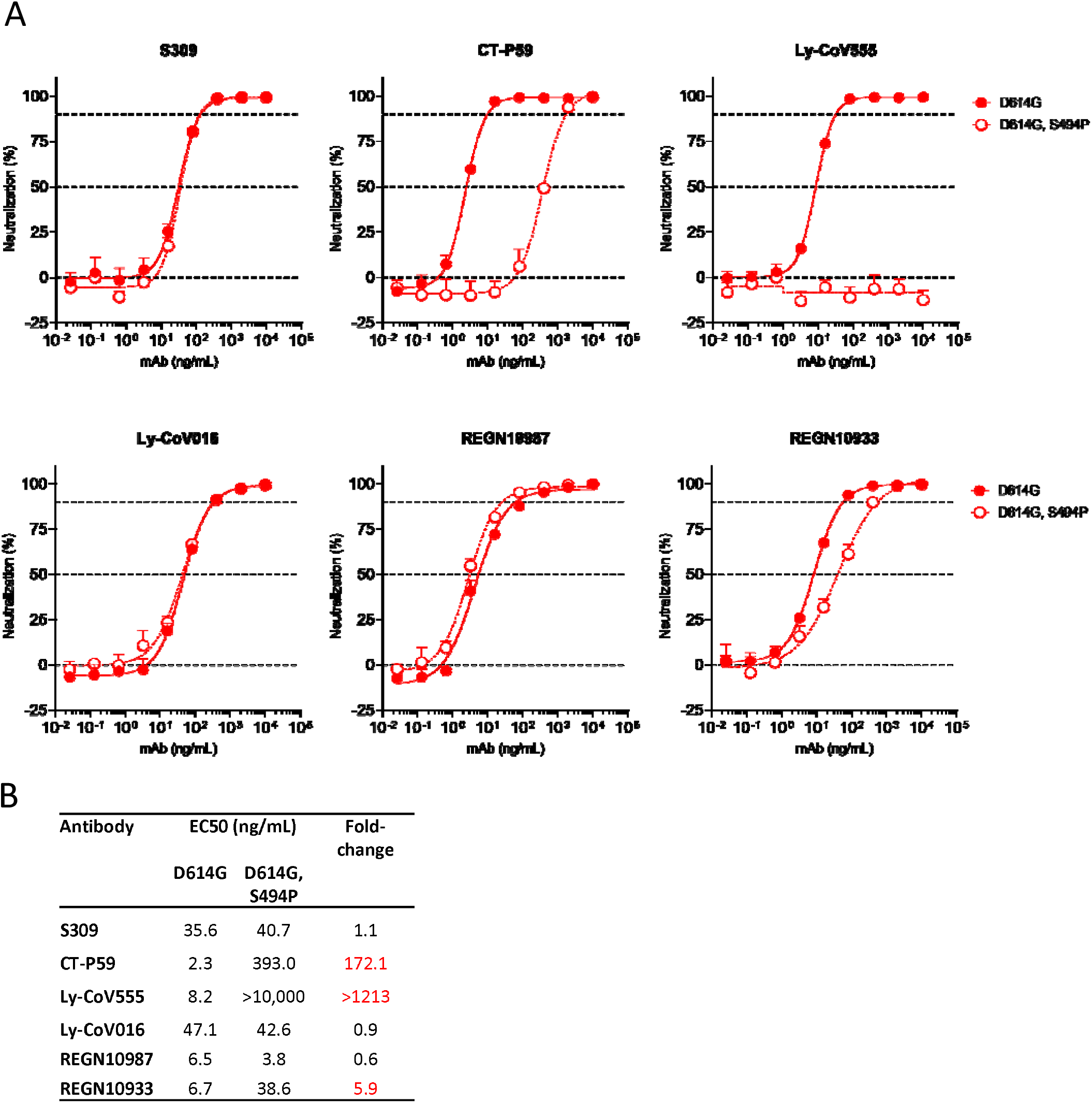
S494P mutation decreases neutralization potential of three clinically-approved therapeutic antibodies. **(A)** VSV-SARS-CoV-2 pseudovirus was generated based on the “Wuhan-Hu-1” sequence with either the D614G mutation or D614G + S494P mutations. Virus neutralization was measured in a microneutralization assay on Vero E6 cells. Exemplary results from one repeat are shown. **(B)** EC50 values and fold-changes were calculated from two independent experiments. S309 is the parent molecule of VIR-7831, which had been previously evaluated on the S494P variant and showed no significant change in neutralization in Cathcart et al^30^.

In summary, we established a method for forecasting spreading mutations and applied it to forecast future contributors to putative VOCs/VOIs. These predictions are consistent with mutations known to be important from *in vitro* data. We find that a subset of forecast mutations could have implications for the continued efficacy of clinical antibodies, but that the level of these concerns varies widely.

## Discussion

We established a working definition for spreading mutations and leveraged this definition to deliver a systematic analysis of amino acid features predictive of mutation spread. This yielded a simple, explainable, and accurate model for forecasting mutations several months in advance, across multiple pandemic waves. Although this model requires nothing more than genomic surveillance data, we also highlight the value of the complete mapping of epitopes, *in vitro* deep site-directed mutagenesis, and downstream functional readouts. Confidence in the prediction of spreading mutations comes through retrospectively evaluating multiple waves of the pandemic and verifying consistency experimental data, and with a plausible causal framework. Furthermore, long observed lags between the earliest warning signals and high population frequency of current mutations in VOC and VOI gives support for using forecasting to anticipate the spread of future concerning mutations. Although this approach will be limited in its ability to anticipate mutations that appear and rise to high frequencies within a short time frame, we found this to be a rare occurrence.

We provide a forecast of the mutations that are most likely to spread over the coming months, and we demonstrate how they could differentially impact clinical antibodies. We intend for these results to provide a foundation for future improvement. Although we have shown that Epi Score is robust to shifting evolutionary dynamics, performance can be monitored in real-time, and if necessary, re-tuned to capture novel behavior. This approach can also be generalized and improved upon to stay ahead of evolutionary cycles for other pathogens^34^, when sufficiently rich and representative genomic sampling is available.

## Data Availability

We gratefully acknowledge the authors, originating and submitting laboratories of the sequences from GISAID used in the current study. The full acknowledgement list can be found in Suppl. File S4.

https://www.gisaid.org/

## Acknowledgments

We gratefully acknowledge the authors, originating and submitting laboratories of the sequences from GISAID used in the current study. The full acknowledgement list can be found in **Suppl. File S4**. We thank Darren Martin and Emma Hodcroft for useful discussion, D.L.R. is funded by the MRC (MC_UU_12014/12).

## Supplemental materials

### Methods

#### Variable definitions and data sources

The definitions of the variables presented, how they are grouped into categories, and where they can be retrieved, can be found in **Suppl. File 1**.

#### Code availability and environment

Analyses on GISAID data extracts were conducted in python (Python Software Foundation. Python Language Reference, version 3.7. Available at http://www.python.org). Code was built in Jupyter lab notebooks^35^ and relied upon a number of common data analysis libraries^36–39^. Code will be made available prior to print publication of this manuscript.

#### Sequence access and alignment

The viral sequences and metadata were obtained from GISAID EpiCoV project (https://www.gisaid.org/). Analysis was performed on sequences submitted to GISAID up to April 24th, 2021. The spike protein sequences were either obtained directly from the protein data file provided by GISAID or, for the latest submitted sequences that were not incorporated yet in the protein data file at the day of data retrieval, from the genomic sequences, with Exonerate ^40^ 2.4.0--haf93ef1_3 (https://quay.io/repository/biocontainers/exonerate?tab=tags) using protein to DNA alignment with parameters -m protein2dna --refine full --minintron 999999 --percent 20 and using accession YP_009724390.1 as a reference.

Multiple sequence alignment of all human spike proteins was performed with mafft^41^ 7.475-- h516909a_0 (https://quay.io/repository/biocontainers/mafft?tab=tags) with parameters –auto -- mapout --reorder --keeplength --addfragments using the same reference as above. Spike sequences that contained >10% ambiguous amino acid or that were less than 80% of the canonical protein length were discarded.

A total of 1,104,875 sequences were used for analysis. Mutations were then extracted as compared to the reference with R 4.0.2 (https://www.r-project.org/) using Biostrings 2.56.0 (https://bioconductor.org/packages/Biostrings) and haplotypes were obtained by combining all amino acid mutations (substitutions, insertions, and deletions) identified on the Spike protein when compared to the reference sequence. We note that GISAID reports viral consensus sequences for individuals. Thus, for our analysis we assume the presence of a single virus in a single sample. This approach will not detect evolution of viral quasi-species within individuals but allows for the characterization of dominant spreading mutations in populations^33^.

#### Defining spreading mutations

As described in the main text, mutations were selected based on a Fisher’s exact test for frequency fold change per country, adjusted for multiple comparisons. This approach was selected after exploratory analysis found that more granular trend tests such as the Mann-Kendall trend test were less favorably powered in low-data countries. Comparisons were adjusted using the function *statsmodels*.*stats*.*multitest*.*fdrcorrection* from the *statsmodels* package^38^ with an alpha of 0.05. This applies a Benjamini-Hochberg correction. We constructed the 2×2 tables used for the Fisher’s exact test in the following manner. Within each country, we tabulated four counts: the number of sequences containing the mutation of interest, versus those that did not; in one window before, and one after the date cutoff (e.g. Nov. 1^st^). From each table, we calculated a fold change and an associated comparison-adjusted p-value. Mutations with a significant p-value from any country were accepted. The number of comparisons for the adjustment was taken as the number of countries times the number of observed mutations worldwide.

#### Analyzed features

We investigated B cell epitopes and CD4+ and CD8+ T epitopes in the viral Spike protein^16,42^ as features that might predict spreading mutations. We also integrated *in vitro* mutagenesis data quantifying ACE2 binding of the viral Spike protein, expression of the viral spike protein, and escape from monoclonal antibody neutralization as measured in pseudovirus assays and/or binding of monoclonal antibodies to the Spike protein^14,21^. In addition, we examined features of viral genome conservation such as RNA secondary structure constraint^22^ and conservation of amino acids, as quantified by Shannon entropy, across the three sarbecovirus clades that encompass both SARS and SARS-CoV-2. We also assessed metrics of positive selection via MEME and FEL^23^. We looked at variation in the viral proteome as captured by novel natural language learning tools^19^. We also evaluated epidemiologic features calculated from the training periods, such as mutation frequency, and the distribution of mutations across countries and viral variant backgrounds. We further calculated an integrated epidemiology score (“Epi Score”) as the exponentially weighted mean ranking across mutation frequency, the fraction of unique variant sequences that contain an amino acid mutation, and the number of countries in which a mutation was been observed. Briefly, to calculate Epi score, we calculated the percentile of each component score (p), and from this calculated a new score (10 ** p). The average of these scores between the metric pair resulted in the combined score that ranged between 1 and 10.

#### Preparing feature sets

Deep mutational scan data^21,43^, were retrieved from the following repository: https://github.com/brianhie/viral-mutation. For T-cell data where scores are associated to oligonucleotides instead of mutations or sites, overlapping scores were averaged per site. When there were multiple experimental conditions, the maximum value per site was taken.

Antibody binding energies were calculated using Molecular modeling software MOE^27^ (v2019.0102). To produce the antibody binding score, we first calculated pairwise binding energies (the sum of van der Waals, ionic, aromatic, and hydrogen-bond interactions) between each residue in the antigen epitope and each residue in the corresponding antibody Fab paratope, including all residues within a cutoff distance of 5.0 Å from the epitope/paratope interface. All structures were prepared prior to these calculations using the structure preparation, protonation and energy minimization steps in MOE, with default settings. The binding energies of each epitope residue that interacted with multiple Fab residues were added together and the percentage of the binding energy contributed by each epitope residue to the total binding energy was calculated.

When more than one copy of the complex was present in the asymmetric unit, binding energy contributions were averaged across all copies. An overall binding energy per site was calculated as the max score across all antibodies.

Interspecies conservation was calculated from a nucleotide multiple sequence alignment of 44 sarbecoviruses. The Shannon entropy was calculated for each column in the alignment. Non-ATGC letters (including gaps) were ignored. RNA structure SHAPE-seq intensities are downloaded from: http://incarnatolab.com/downloads/datasets/SARS_Manfredonia_2020/XML.tar.gz)^44^. They are post processed by taking the mean of each 12-nucleotide sliding window and the window centered on a given nucleotide is used.

For natural language processing (NLP) neural network features, we used the grammaticality and semantic change scores reported by Hie et al.^18^ in which a bidirectional long short-term memory (BiLSTM) model was trained on Spike sequences from GISAID and GenBank. We obtained two versions of the model: the original model trained on sequences through sampled prior to June 1, 2020, and a second model that was retrained starting from random weight initializations on GISAID Spike sequences sampled prior to November 1, 2020. For all prediction periods after November 1st, the latter model was used. For prediction periods before this time, the former model was employed.

Natural selection features were generated using MEME^45^ and FEL^23^ methods implemented in the HyPhy package^24^ (version 2.5.31). Data preparation, alignment, and tree inference were performed using an existing pipeline (https://github.com/veg/SARS-CoV-2/tree/compact/). Briefly, the pipeline curates sequences to remove low quality genomes and filter out potential sequencing errors and compresses the input to unique haplotypes over each gene region. A codon-aware mapping and multiple sequence alignment of gene regions is followed by rapid phylogenetic tree inference, and site-level selection analyses applied to internal tree branches (a standard procedure for viral intra-species data). FEL tests for pervasive negative or positive selection, while MEME tests for episodic positive selection. Both tests report p-values (based on the likelihood ratio tests); MEME further reports the number of branches which provide support for the positive selection model component.

For epidemiologic variables, the “fraction of unique haplotypes” metric was defined as the proportion of the known haplotype backgrounds in which a given mutation occurred. The “mutation frequency” metric is defined as the fraction of sequenced individuals who had a mutation at that site. The “number of countries” metric is defined as the number of countries in which a mutation is observed in at least two sequences.

Epi Score was calculated as an exponentially weighted mean of the mutation ranks according to mutation frequency, fraction of unique haplotypes in which the mutation occurs, and the number of countries in which it occurs. Specifically, this involved (i) calculating the percentile for each score, for each metric, (ii) exponentiating percentile to the power of 10, and (iii) averaging these exponentiated percentiles. The effect of this procedure is to assign highly differentiated weights to high rankings, and relatively small and similar weights to mutations that are not at the top of the list. For example, top ranked mutation versus a 90^th^ percentile mutation will have a score difference of 2.1 (10 vs 7.9), whereas a mutation at the 50^th^ percentile and one at the 40^th^percentile will have a score difference of 0.65 (3.16 vs 2.51). This scheme is particularly advantageous if measurements for lower-ranked entities are more noisy than higher ranked ones, and/or if one wants to up-weight high rankings.

For conversion from mutation-to site-level scores, the site level score was taken to be the maximum of mutation scores at that position. For the conversion of site-to mutation-level scores, the site-level score was assigned to all observed mutation at that position. In cases where data needed to be imputed, min-imputation was performed. For example, all sites without measured antibody binding energies were assigned a binding energy of zero. For all metrics, in cases of multiple experimental conditions, the max score per site or mutation (as appropriate) was taken. This was appropriate because the few metrics where lower scores implied a higher probability of spread (e.g. MEME p-values) did not have missing values.

#### Quantifying predictive performance

Predictive performance was quantified using the area under the receiver operator characteristic curve (AUROC). This quantity can be interpreted as the probability that a given score correctly ranks a random pair of positive and negative examples. Performance was assessed by two methods: (i) direct univariate ranking and (ii) model fitting with sets of features. The AUROC for univariate ranking was calculated as the maximum AUROC upon sorting by that metric in either ascending or descending order. Receiver Operator Characteristic (ROC) curves were generated by varying the numerical cutoff ‘C’ on each metric beyond which a mutation is called to be spreading. Given these calls, we then calculated sensitivity and specificity values for each value of C tested. Plotting sensitivity versus specificity yields the ROC curve. The area under the ROC (AUROC) was then used to quantify the capacity for that variable to distinguish spreading from non-spreading amino acid mutations.

For model fitting, performance was assessed by cross-validation. This involves partitioning the data into chunks or “folds” and iteratively predicting each (test) fold based on training with all the other chunks. In this procedure, it is important to make sure that correlated observations are kept in the same fold so that information does not leak between the training folds and the test fold. As a hypothetical example, a model could memorize the attributes of one identical twin in a training set to predict the values of the other in the test set. In the case of mutations, we were concerned that the co-occurrence of mutations on the same haplotypes could introduce a correlation in their metrics. To mitigate this issue, we made sure mutations from the same clade were always included in the same fold. Clades were defined according to GISAID annotation. The following clades were used to define folds: G, GH, GR, GRY. The remaining smaller clades were pooled into a single fold. This resulted in five folds ranging in size from around 700 to 2000 mutations. AUROC values were then calculated within each test fold and averaged across test folds to yield an overall performance.

#### Predictive performance of sets of features

Prediction was performed using forward feature selection followed by logistic regression. The criterion for forward selection was cross-validated AUROC of the logistic regression model within the training set. Feature selection and model fitting were performed separately within each fold of the outer cross validation loop. Logistic regression was chosen due to its sample efficiency. We found that random forest classifiers obtained worse performance. Similarly, we found that combined models did worse than individual features if there was no feature selection step. We also tried a select K best feature selection strategy, which generally recapitulated the performance of the single best feature. We interpret these results to mean that limited sample size amplifies the effect of noisy features, and that greedily selecting for high AUROC features does not do a good job of selecting for complementarity. The members of each of the feature sets are enumerated in **Suppl. File 1**.

#### Selected features

Since a different model is fit for each cross-validation fold, we retrained a single model on all data to produce a single set of selected features for each feature set. The selected features will be presented in our GitHub repository upon peer-reviewed publication of the manuscript.

#### Mediation analysis

The strength of predictions based solely on the epidemiological features led the study to consider a hypothesized causal model (**Fig. 4C**) to explain the effectiveness of these features relative to the contribution of biological measurements. We propose that the biological factors that we analyze determine viral fitness, which in turn drives spread, as measured via epidemiology. As illustrated in (**Fig. 4C**), epidemiology and evolution-based measures both draw on empirical variation, as captured by GISAID. We hypothesized that epidemiologic variables demonstrated superior performance because they are most proximal to the outcome variable, and therefore mediate the effects of the other variables. In causal inference, a mediated variable is a quantity that indirectly contributes to an outcome of interest (in this case spreading mutations) by altering an intermediate factor (a mediator; e.g., initial mutation spread). The classical Baron and Kenny test for mediation can be divided into three steps (i) make sure the variable of interest predicts the outcome, (ii) verify that the variable of interest predicts the mediator, and (iii) show that the variable of interest does not add to the predictive performance of the mediator when including both in a single model.

Step 1 was performed as part of the baseline analysis, and the complete results of this can be found in **Suppl. Figs. S2A and S2B**. For step 2, since few variables showed above-random performance outside of the RBD, we focused our analysis within the RBD. We attempted to predict the putative mediator (Epi Score). We predicted this surrogate outcome by first binarizing it to indicate whether the mutation score was in the top N mutations, where N is two times the number of mutations that spread in the observed dataset. We chose to multiply by two after consulting the positive predictive values in **Suppl. Fig S5**. These results, shown by comparing the first and second columns in **Fig 4C**, demonstrate that variables that are predictive of spread are also predictive of our epidemiologic predictor, with approximately the same magnitude. Therefore, we can conclude that criteria 1 & 2 have been fulfilled.

Finally, we fit models with each variable in addition to the epidemiologic predictor to test for complementarity. Since we are most interested in the RBD due to data availability, we encountered the issue that supervised models trained on full length spike tended to perform poorly with variables that are only observed within the RBD. Specifically, we saw that supervised models significantly decreased in performance when including these variables, indicating overfitting. To address this issue, and to make our results more comparable to the univariate analysis, we generated a single score from the variable pairs by exponentially weighting the ranks of each metric. This was performed according to the same procedure as the Epi Score. Specifically, we calculated the percentile of each score (p), and from this calculated a new score (10 ** p). The average of these scores between the metric pair resulted in the combined score.

#### Testing integrated predictive models across waves and time lags

For testing predictive models across different waves and time lags, below are the time periods that we used for wave 2. The first group denotes the feature calculation window, and the second group of dates in each set denote the time window in which variant growth was assessed.

[(“2020-01”, “2020-02”), (“2020-06”, “2020-07”, “2020-08”)],

[(“2020-01”, “2020-02”, “2020-03”), (“2020-06”, “2020-07”, “2020-08”)],

[(“2020-02”, “2020-03”, “2020-04”), (“2020-06”, “2020-07”, “2020-08”)],

[(“2020-03”, “2020-04”, “2020-05”), (“2020-06”, “2020-07”, “2020-08”)]

Below are the time periods used for wave 3.

[(“2020-05”, “2020-06”, “2020-07”), (“2020-11”, “2020-12”, “2021-01”)],

[(“2020-06”, “2020-07”, “2020-08”), (“2020-11”, “2020-12”, “2021-01”)],

[(“2020-07”, “2020-08”, “2020-09”), (“2020-11”, “2020-12”, “2021-01”)],

[(“2020-08”, “2020-09”, “2020-10”), (“2020-11”, “2020-12”, “2021-01”)]

#### Forecasting spreading mutations

The list of forecast mutations was generated by calculating Epi Score on the most recent three months of data and taking the top 200 ranked mutations. The threshold of 200 mutations was chosen based on the analysis presented in **Suppl. Fig. S5**.

#### Definition of Variants of Concern

Variants of concern were defined as those specified by the CDC, plus additional mutations which occurred at a rate of 80% of the most prevalent variant in the lineage^3^:

Alpha, B.1.1.7 (CDC VOCs): H69-, V70-, Y144-, N501Y, A570D, D614G, P681H, T716I, S982A, D1118H

Beta, B.1.351(CDC VOCs): K417N, E484K, N501Y, D614G

B.1.351 (additional associated mutations: D80A, D215G, L241-, L242-, A243-, A701V Gamma, P.1 (CDC VOCs): K417T, E484K, N501Y, D614G

P.1 (additional associated mutations): L18F, T20N, P26S, D138Y, R190S, H655Y, T1027I, V1176FB.1.427 (CDC VOCs): L452R, D614G

Epsilon, B.1.427 (additional associated mutations): S13I, W152C

B.1.429 (CDC VOCs): S13I, W152C, L452R, D614G

#### SARS-CoV-2 pseudotyped VSV production and neutralization

To generate SARS-CoV-2 pseudotyped vesicular stomatitis virus, Lenti-X 293T cells (Takara) were seeded in 10-cm dishes for 80%. next day confluency. The next day, cells were transfected with a plasmid encoding for SARS-CoV-2 S-glycoprotein (YP_009724390.1) harboring a C-terminal 19 aa truncation and the D614G or D614G + S494P mutations using TransIT-Lenti (Mirus Bio) according to the manufacturer’s instructions. One day post-transfection, cells were infected with VSV(G*ΔG-luciferase) (Kerafast) at an MOI of 3 infectious units/cell. Viral inoculum was washed off after one hour and cells were incubated for another day at 37°C. The cell supernatant containing SARS-CoV-2 pseudotyped VSV was collected at day 2 post-transfection, centrifuged at 1000 x g for 5 minutes to remove cellular debris, aliquoted, and frozen at −80°C.

For viral neutralization, Vero E6 cells were seeded into black-walled, clear-bottom 96-well plates at 20,000 cells/well and cultured overnight at 37°C. The next day, 9-point 5-fold serial dilutions of antibodies were prepared in media. SARS-CoV-2 pseudotyped VSV was diluted 1:20 in media and added 1:1 to each antibody dilution. Virus:antibody mixtures were incubated for 1 hour at 37°C. Media was removed from the cells and 50 μL of virus:antibody mixtures were added to the cells. One hour post-infection, 100 μL of media was added to all wells and incubated for 17-20 hours at 37°C. Media was removed and 50 μL of Bio-Glo reagent (Promega) was added to each well. The plate was shaken on a plate shaker at 300 RPM at room temperature for 15 minutes and RLUs were read on an EnSight plate reader (Perkin-Elmer).

## References

1. McCallum, M. et al. N-terminal domain antigenic mapping reveals a site of vulnerability for SARS-CoV-2. Cell (2021) doi:10.1016/j.cell.2021.03.028.

2. Elbe, S. & Buckland-Merrett, G. Data, disease and diplomacy: GISAID’s innovative contribution to global health. Global Challenges 1, 33–46 (2017).

3. Control, C. for D. SARS-CoV-2 Variants of Concern. https://www.cdc.gov/coronavirus/2019-ncov/cases-updates/variant-surveillance/variant-info.html (n.d.).

4. Adiga, A. et al. All Models Are Useful: Bayesian Ensembling for Robust High Resolution COVID-19 Forecasting. Medrxiv 2021.03.12.21253495 (2021) doi:10.1101/2021.03.12.21253495.

5. Zhao, H. et al. COVID-19: Short term prediction model using daily incidence data. Plos One 16, e0250110 (2021).

6. Ray, E. L. et al. Ensemble Forecasts of Coronavirus Disease 2019 (COVID-19) in the U.S. Medrxiv 2020.08.19.20177493 (2020) doi:10.1101/2020.08.19.20177493.

7. Control, C. for D. COVID-19 Forecasts: Cases. https://www.cdc.gov/coronavirus/2019-ncov/cases-updates/forecasts-cases.html (n.d.).

8. Padane, A. et al. First detection of the British variant of SARS-CoV-2 in Senegal. New Microbes New Infect 100877 (2021) doi:10.1016/j.nmni.2021.100877.

9. Valesano, A. L. et al. Temporal dynamics of SARS-CoV-2 mutation accumulation within and across infected hosts. Plos Pathog 17, e1009499 (2021).

10. Charkiewicz, R. et al. The first SARS-CoV-2 genetic variants of concern (VOC) in Poland: The concept of a comprehensive approach to monitoring and surveillance of emerging variants. Adv Med Sci 66, 237–245 (2021).

11. Dejnirattisai, W. et al. Antibody evasion by the P.1 strain of SARS-CoV-2. Cell (2021) doi:10.1016/j.cell.2021.03.055.

12. Collier, D. A. et al. Sensitivity of SARS-CoV-2 B.1.1.7 to mRNA vaccine-elicited antibodies. Nature 1–10 (2021) doi:10.1038/s41586-021-03412-7.

13. Starr, T. N., Greaney, A. J., Dingens, A. S. & Bloom, J. D. Complete map of SARS-CoV-2 RBD mutations that escape the monoclonal antibody LY-CoV555 and its cocktail with LY-CoV016. Cell Reports Medicine 100255 (2021) doi:10.1016/j.xcrm.2021.100255.

14. Starr, T. N. et al. Deep mutational scanning of SARS-CoV-2 receptor binding domain reveals constraints on folding and ACE2 binding. Cell (2020) doi:10.1016/j.cell.2020.08.012.

15. Starr, T. N. et al. Prospective mapping of viral mutations that escape antibodies used to treat COVID-19. Science 371, 850–854 (2021).

16. Agerer, B. et al. SARS-CoV-2 mutations in MHC-I-restricted epitopes evade CD8+ T cell responses. Sci Immunol 6, eabg6461 (2021).

17. Tarke, A. et al. Negligible impact of SARS-CoV-2 variants on CD4+ and CD8+ T cell reactivity in COVID-19 exposed donors and vaccinees. Biorxiv 2021.02.27.433180 (2021) doi:10.1101/2021.02.27.433180.

18. Hie, B., Zhong, E. D., Berger, B. & Bryson, B. Learning the language of viral evolution and escape. Science 371, 284–288 (2021).

19. Weisblum, Y. et al. Escape from neutralizing antibodies by SARS-CoV-2 spike protein variants. Elife 9, e61312 (2020).

20. Hoffmann, M., Kleine-Weber, H. & Pöhlmann, S. A Multibasic Cleavage Site in the Spike Protein of SARS-CoV-2 Is Essential for Infection of Human Lung Cells. Mol Cell 78, 779–784.e5 (2020).

21. Greaney, A. J. et al. Complete Mapping of Mutations to the SARS-CoV-2 Spike Receptor-Binding Domain that Escape Antibody Recognition. Cell Host Microbe 29, 44-57.e9 (2021).

22. Hodcroft, E. B. et al. Emergence and spread of a SARS-CoV-2 variant through Europe in the summer of 2020. Medrxiv 2020.10.25.20219063 (2021) doi:10.1101/2020.10.25.20219063.

23. Pond, S. L. K. & Frost, S. D. W. Not So Different After All: A Comparison of Methods for Detecting Amino Acid Sites Under Selection. Mol Biol Evol 22, 1208–1222 (2005).

24. Pond, S. L. K. et al. HyPhy 2.5—A Customizable Platform for Evolutionary Hypothesis Testing Using Phylogenies. Mol Biol Evol 37, 295–299 (2019).

25. Martin, D. P. et al. The emergence and ongoing convergent evolution of the N501Y lineages coincides with a major global shift in the SARS-CoV-2 selective landscape. Medrxiv 2021.02.23.21252268 (2021) doi:10.1101/2021.02.23.21252268.

26. Faria, N. R. et al. Genomics and epidemiology of the P.1 SARS-CoV-2 lineage in Manaus, Brazil. Science eabh2644 (2021) doi:10.1126/science.abh2644.

27. Vilar, S., Cozza, G. & Moro, S. Medicinal Chemistry and the Molecular Operating Environment (MOE): Application of QSAR and Molecular Docking to Drug Discovery. Curr Top Med Chem 8, 1555–1572 (2008).

28. Maher, M. C., Uricchio, L. H., Torgerson, D. G. & Hernandez, R. D. Population Genetics of Rare Variants and Complex Diseases. Hum Hered 74, 118–128 (2013).

29. Pearce, N. & Lawlor, D. A. Causal inference—so much more than statistics. Int J Epidemiol 45, 1895–1903 (2016).

30. Cathcart, A. L. et al. The dual function monoclonal antibodies VIR-7831 and VIR-7832 demonstrate potent in vitro and in vivo activity against SARS-CoV-2. Biorxiv 2021.03.09.434607 (2021) doi:10.1101/2021.03.09.434607.

31. Chakraborty, S. Evolutionary and structural analysis elucidates mutations on SARS-CoV2 spike protein with altered human ACE2 binding affinity. Biochem Bioph Res Co 534, 374–380 (2020).

32. McCallum, M. et al. SARS-CoV-2 immune evasion by variant B.1.427/B.1.429. bioRxiv (2021) doi:10.1101/2021.03.31.437925.

33. Peacock, T. P. et al. The furin cleavage site in the SARS-CoV-2 spike protein is required for transmission in ferrets. Nat Microbiol 1–11 (2021) doi:10.1038/s41564-021-00908-w.

34. Tang, J. W. et al. Global epidemiology of non-influenza RNA respiratory viruses: data gaps and a growing need for surveillance. Lancet Infect Dis 17, e320–e326 (2017).

35. Kluyver, T. et al. Jupyter Notebooks – a publishing format for reproducible computational workflows ePrints Soton. in 20th International Conference on Electronic Publishing (2016).

36. Harris, C. R. et al. Array programming with NumPy. Nature 585, 357–362 (2020).

37. Pedregosa, F. et al. Scikit-learn: Machine Learning in Python. Journal of Machine Learning Research 12, 2825–2830 (2011).

38. Seabold, S. & Perktold, J. statsmodels: Econometric and statistical modeling with python. in 9th Python in Science Conference (2010).

39. McKinney, W. Data Structures For Statistical Computing in Python. in Proceedings of the 9th Python Science Conference (eds. Walt, S. van der & Millman, J.) 56–61 (2010). doi:10.25080/majora-92bf1922-00a.

40. Slater, G. S. C. & Birney, E. Automated generation of heuristics for biological sequence comparison. Bmc Bioinformatics 6, 31 (2005).

41. Katoh, K. & Standley, D. M. MAFFT Multiple Sequence Alignment Software Version 7: Improvements in Performance and Usability. Mol Biol Evol 30, 772–780 (2013).

42. Tarke, A. et al. Comprehensive analysis of T cell immunodominance and immunoprevalence of SARS-CoV-2 epitopes in COVID-19 cases. Cell Reports Medicine 2, 100204 (2021).

43. Greaney, A. J. et al. Comprehensive mapping of mutations in the SARS-CoV-2 receptor-binding domain that affect recognition by polyclonal human plasma antibodies. Cell Host Microbe 29, 463–476.e6 (2021).

44. Manfredonia, I. et al. Genome-wide mapping of therapeutically-relevant SARS-CoV-2 RNA structures. Biorxiv 2020.06.15.151647 (2020) doi:10.1101/2020.06.15.151647.

45. Murrell, B. et al. Detecting Individual Sites Subject to Episodic Diversifying Selection. Plos Genet 8, e1002764 (2012).

