## Supplemental Figures and Tables for "Predicting the mutational drivers of future SARS-CoV-2 variants of concern"

### Slide 1
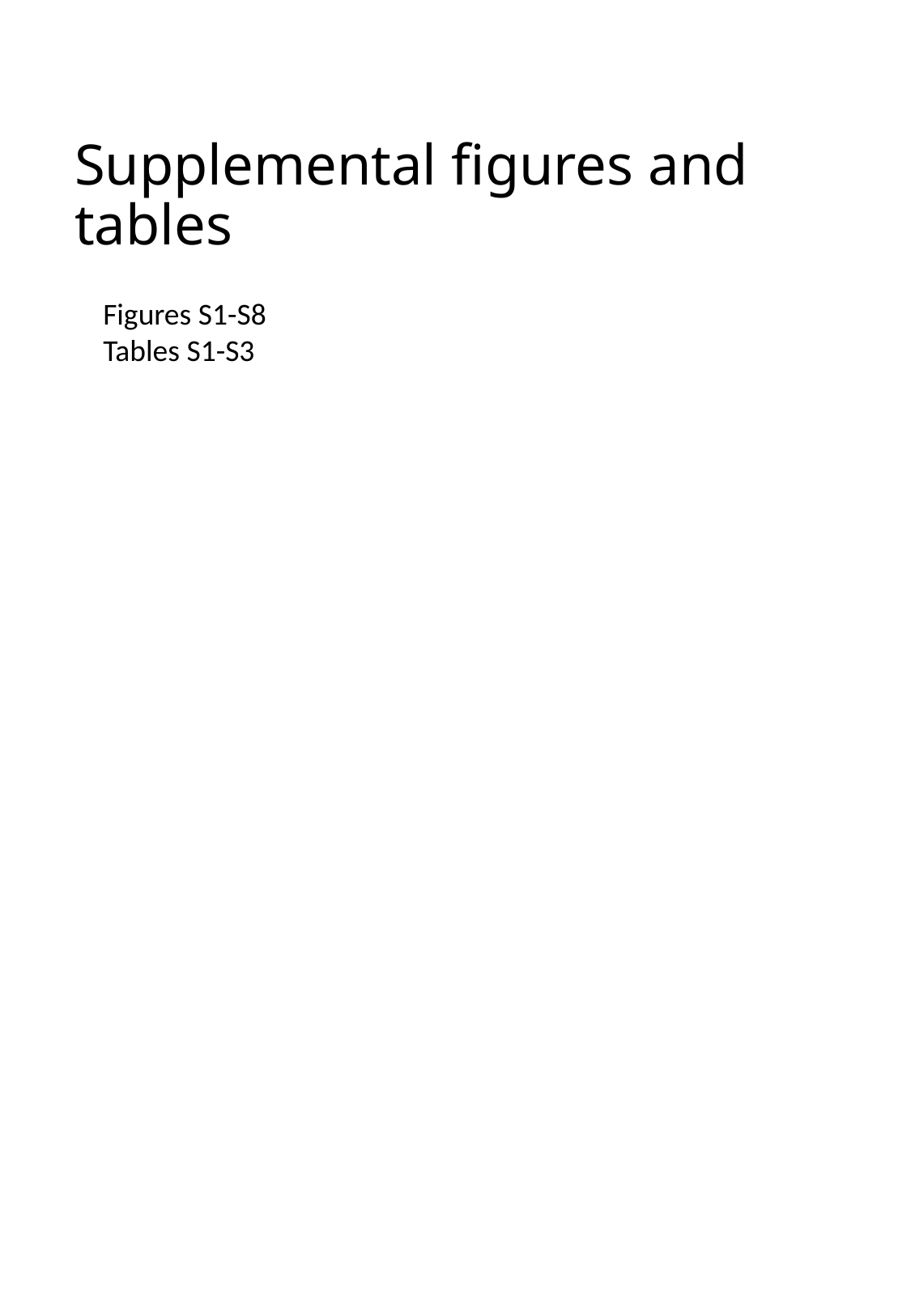

#### Supplemental figures and tables
Figures S1-S8
Tables S1-S3

### Slide 2
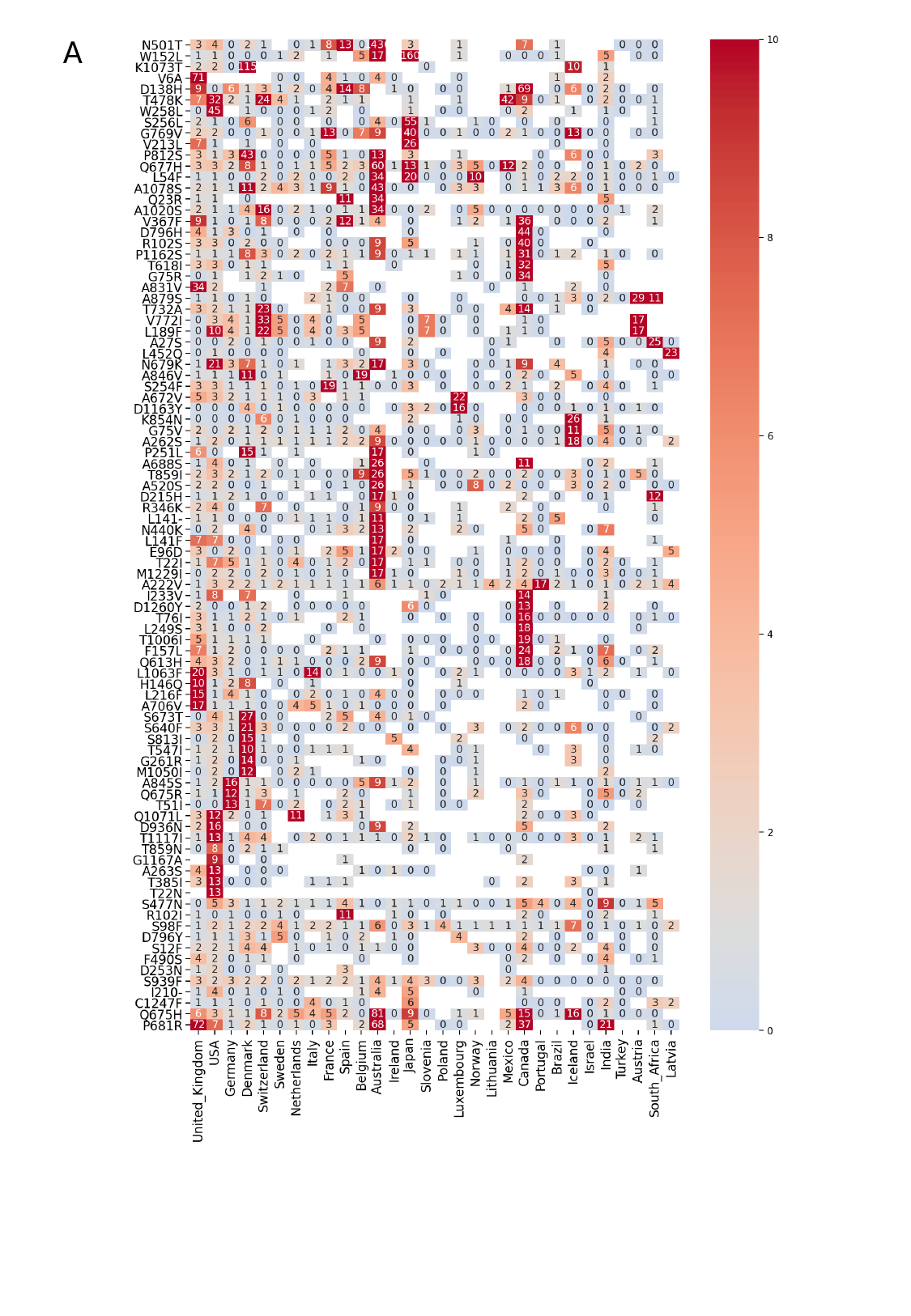

A

### Slide 3
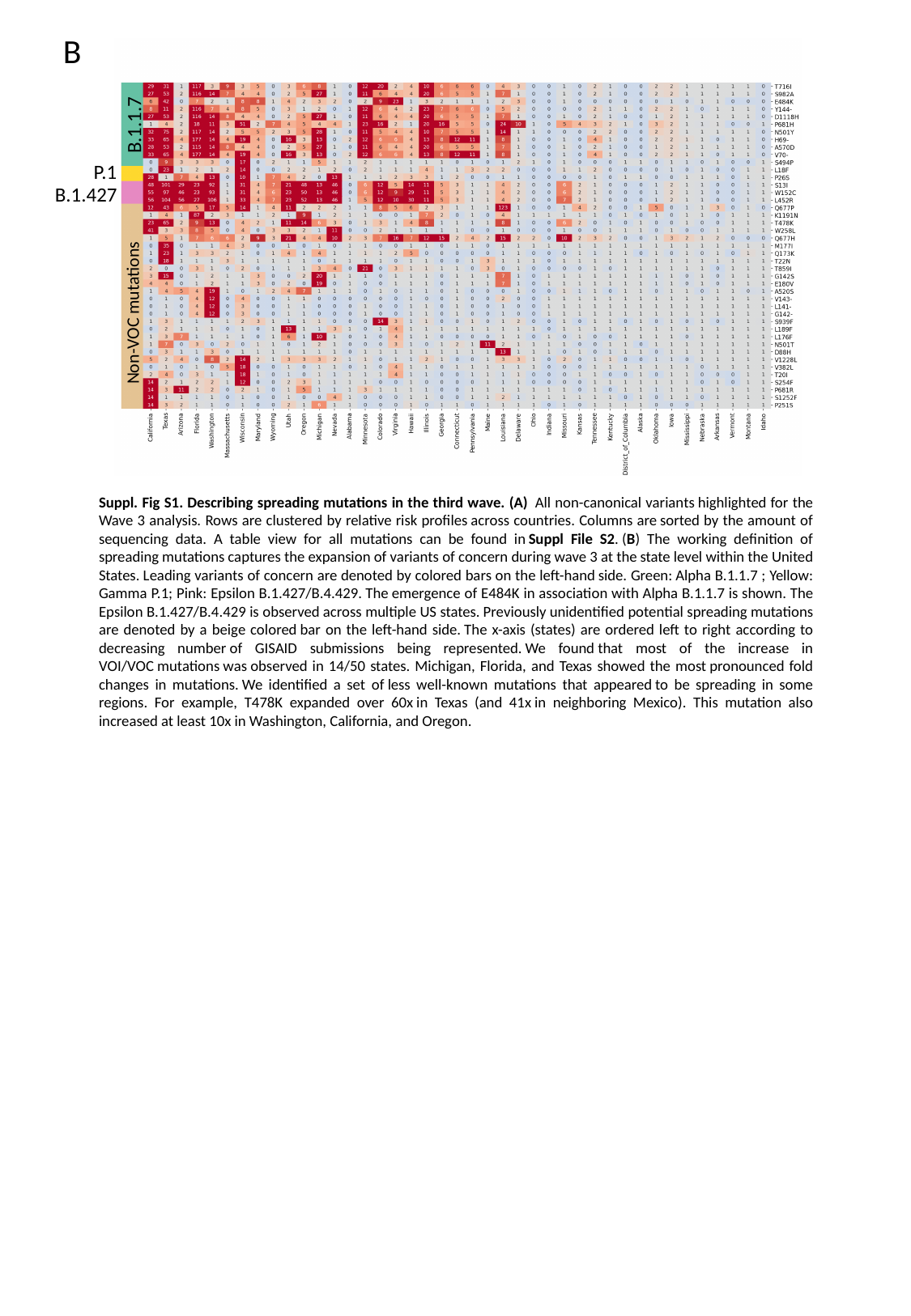

B
B.1.1.7
P.1
B.1.427
Non-VOC mutations
Suppl. Fig S1. Describing spreading mutations in the third wave. (A)  All non-canonical variants highlighted for the Wave 3 analysis. Rows are clustered by relative risk profiles across countries. Columns are sorted by the amount of sequencing data. A table view for all mutations can be found in Suppl File S2. (B) The working definition of spreading mutations captures the expansion of variants of concern during wave 3 at the state level within the United States. Leading variants of concern are denoted by colored bars on the left-hand side. Green: Alpha B.1.1.7 ; Yellow: Gamma P.1; Pink: Epsilon B.1.427/B.4.429. The emergence of E484K in association with Alpha B.1.1.7 is shown. The Epsilon B.1.427/B.4.429 is observed across multiple US states. Previously unidentified potential spreading mutations are denoted by a beige colored bar on the left-hand side. The x-axis (states) are ordered left to right according to decreasing number of GISAID submissions being represented. We found that most of the increase in VOI/VOC mutations was observed in 14/50 states. Michigan, Florida, and Texas showed the most pronounced fold changes in mutations. We identified a set of less well-known mutations that appeared to be spreading in some regions. For example, T478K expanded over 60x in Texas (and 41x in neighboring Mexico). This mutation also increased at least 10x in Washington, California, and Oregon.

### Slide 4
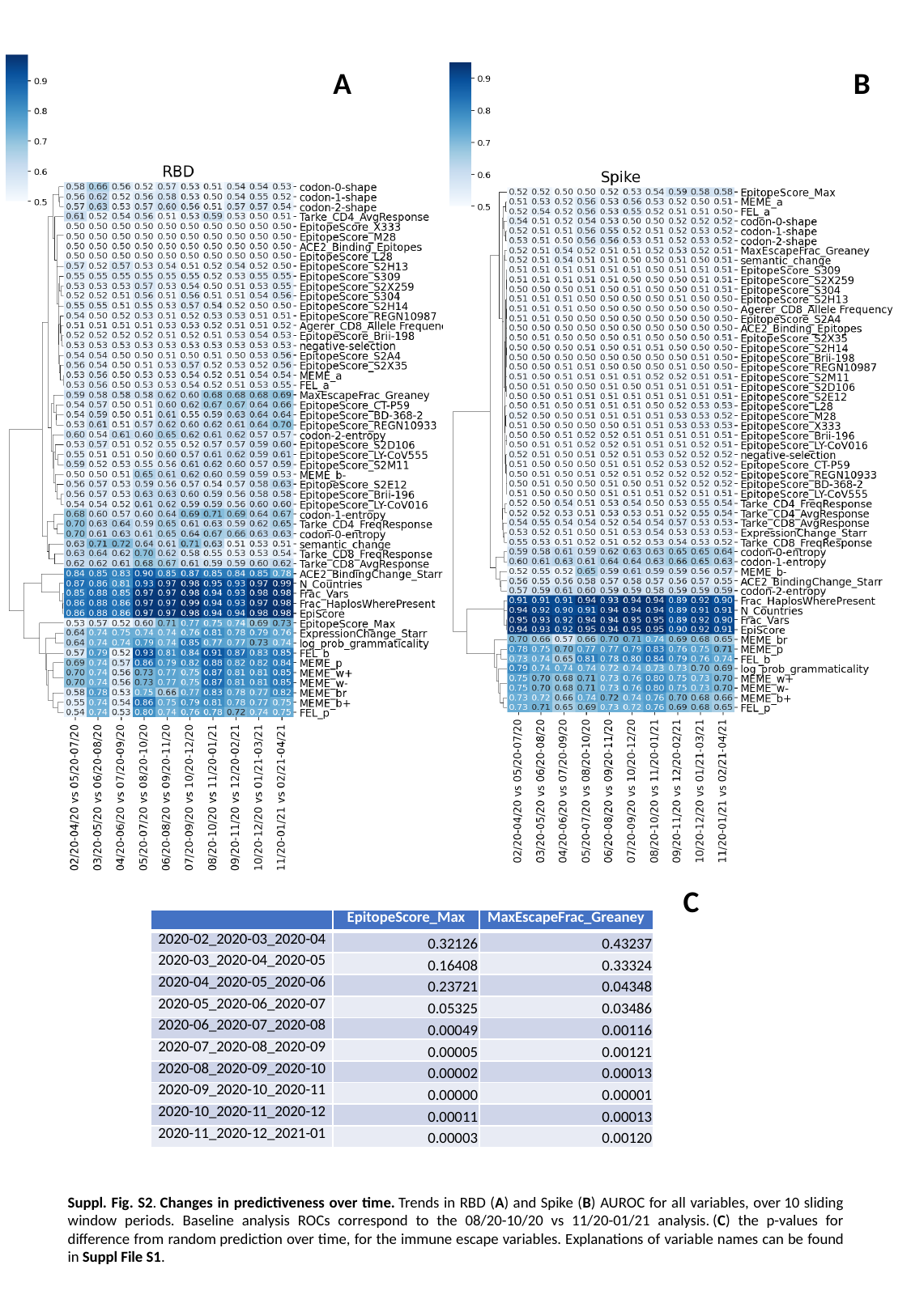

A
B
C
| | EpitopeScore\_Max | MaxEscapeFrac\_Greaney |
| --- | --- | --- |
| 2020-02\_2020-03\_2020-04 | 0.32126 | 0.43237 |
| 2020-03\_2020-04\_2020-05 | 0.16408 | 0.33324 |
| 2020-04\_2020-05\_2020-06 | 0.23721 | 0.04348 |
| 2020-05\_2020-06\_2020-07 | 0.05325 | 0.03486 |
| 2020-06\_2020-07\_2020-08 | 0.00049 | 0.00116 |
| 2020-07\_2020-08\_2020-09 | 0.00005 | 0.00121 |
| 2020-08\_2020-09\_2020-10 | 0.00002 | 0.00013 |
| 2020-09\_2020-10\_2020-11 | 0.00000 | 0.00001 |
| 2020-10\_2020-11\_2020-12 | 0.00011 | 0.00013 |
| 2020-11\_2020-12\_2021-01 | 0.00003 | 0.00120 |
Suppl. Fig. S2. Changes in predictiveness over time. Trends in RBD (A) and Spike (B) AUROC for all variables, over 10 sliding window periods. Baseline analysis ROCs correspond to the 08/20-10/20 vs 11/20-01/21 analysis. (C) the p-values for difference from random prediction over time, for the immune escape variables. Explanations of variable names can be found in Suppl File S1.

### Slide 5
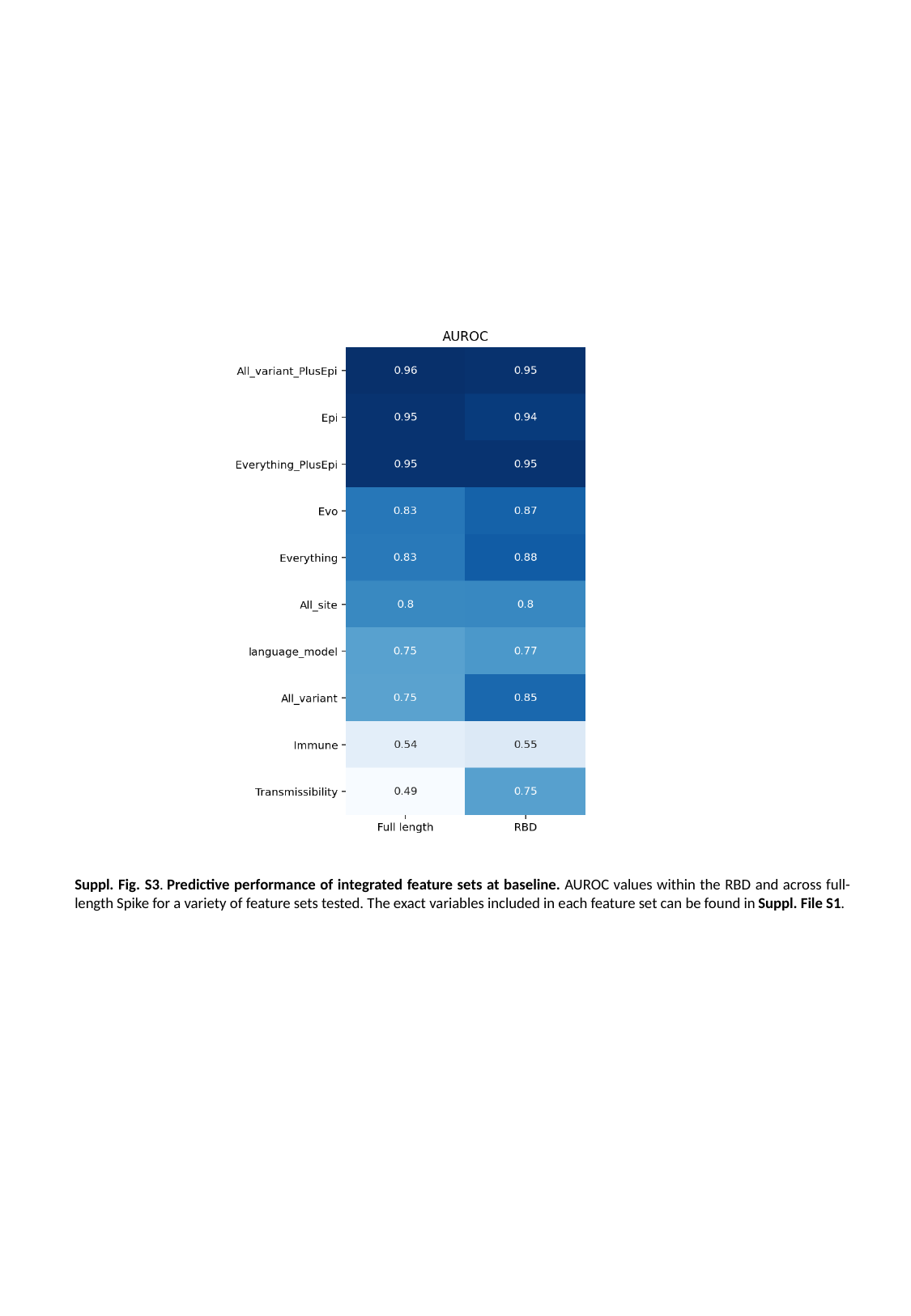

Suppl. Fig. S3. Predictive performance of integrated feature sets at baseline. AUROC values within the RBD and across full-length Spike for a variety of feature sets tested. The exact variables included in each feature set can be found in Suppl. File S1.

### Slide 6
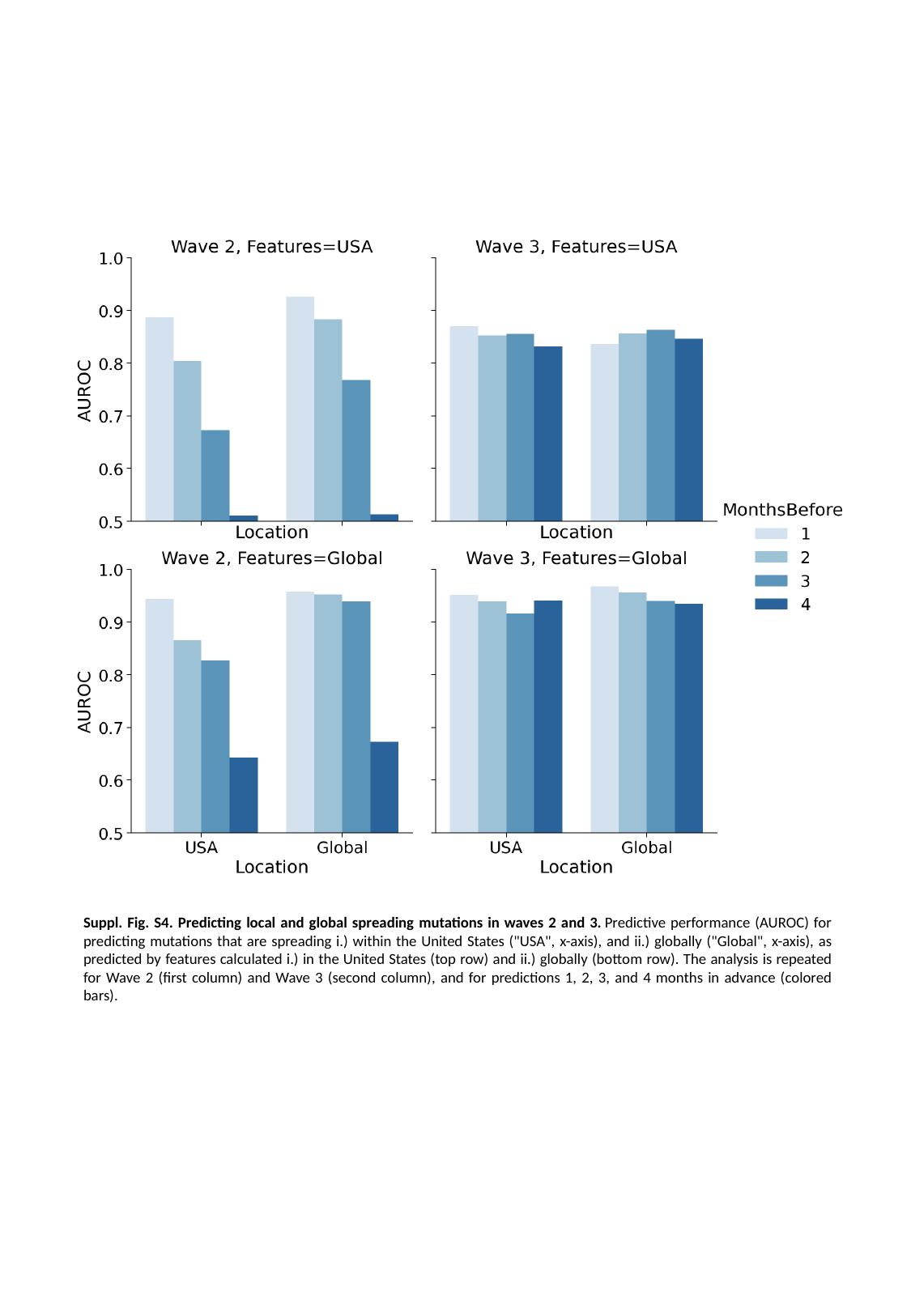

Suppl. Fig. S4. Predicting local and global spreading mutations in waves 2 and 3. Predictive performance (AUROC) for predicting mutations that are spreading i.) within the United States ("USA", x-axis), and ii.) globally ("Global", x-axis), as predicted by features calculated i.) in the United States (top row) and ii.) globally (bottom row). The analysis is repeated for Wave 2 (first column) and Wave 3 (second column), and for predictions 1, 2, 3, and 4 months in advance (colored bars).

### Slide 7
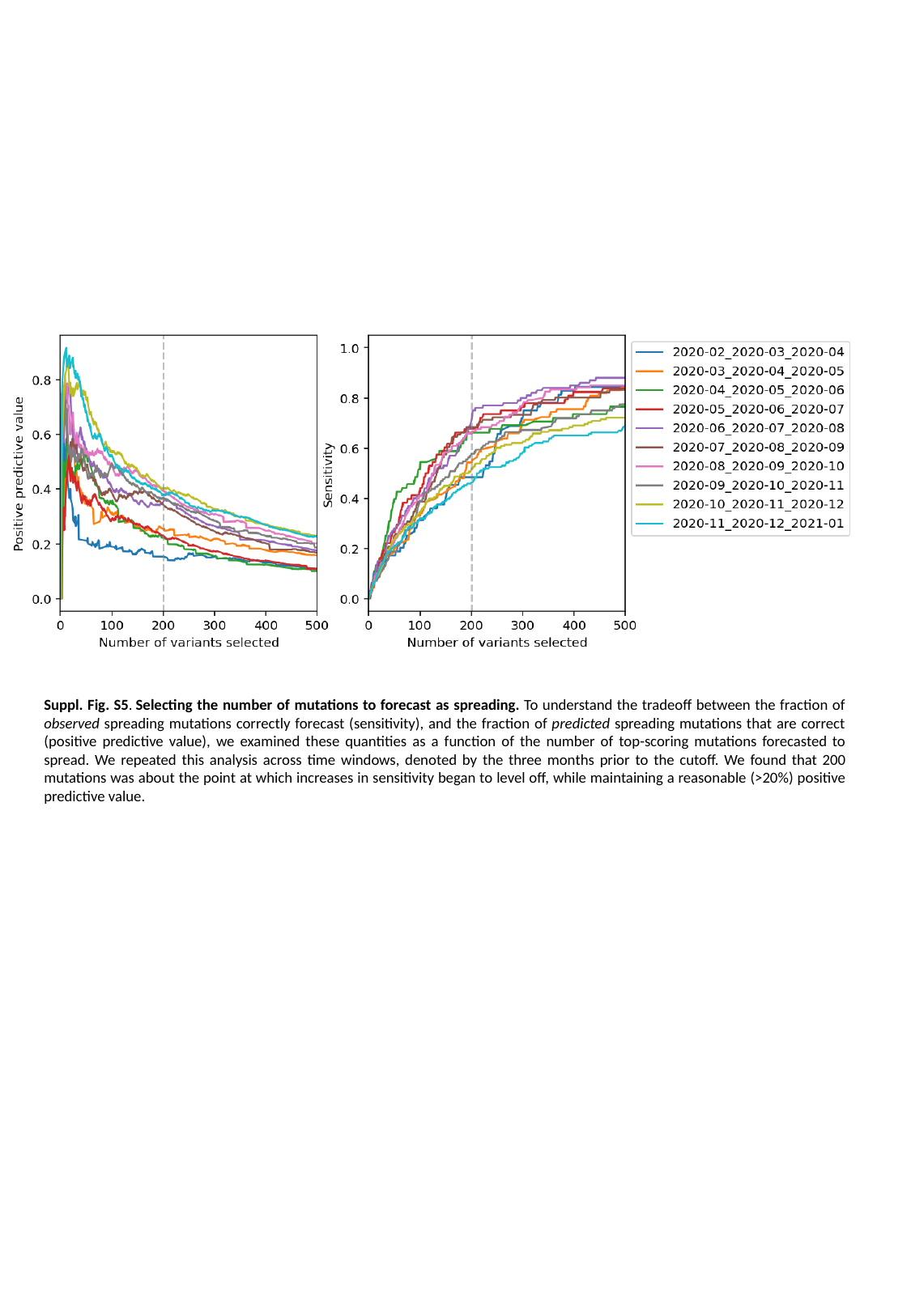

Suppl. Fig. S5. Selecting the number of mutations to forecast as spreading. To understand the tradeoff between the fraction of observed spreading mutations correctly forecast (sensitivity), and the fraction of predicted spreading mutations that are correct (positive predictive value), we examined these quantities as a function of the number of top-scoring mutations forecasted to spread. We repeated this analysis across time windows, denoted by the three months prior to the cutoff. We found that 200 mutations was about the point at which increases in sensitivity began to level off, while maintaining a reasonable (>20%) positive predictive value.

### Slide 8
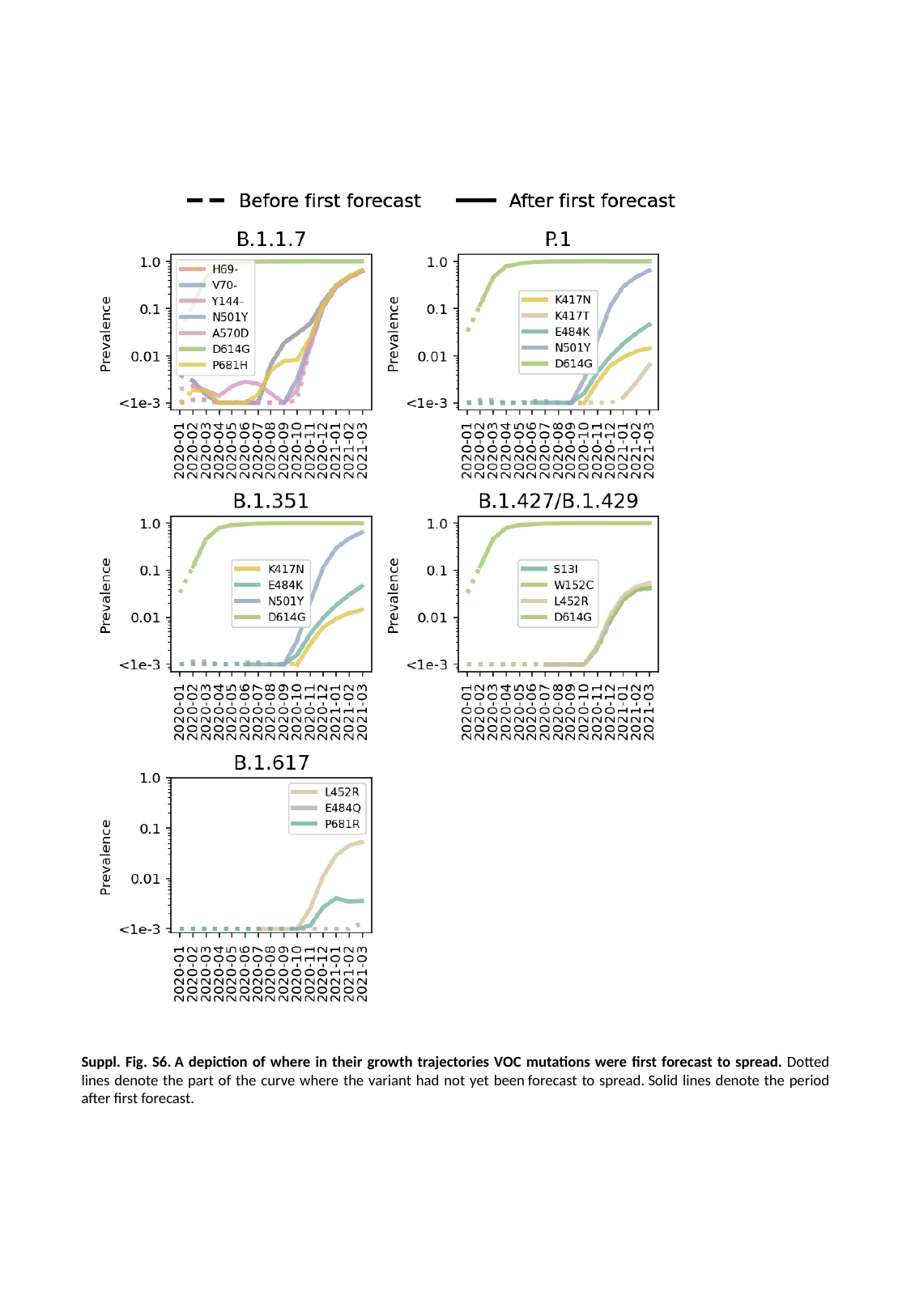

Suppl. Fig. S6. A depiction of where in their growth trajectories VOC mutations were first forecast to spread. Dotted lines denote the part of the curve where the variant had not yet been forecast to spread. Solid lines denote the period after first forecast.

### Slide 9
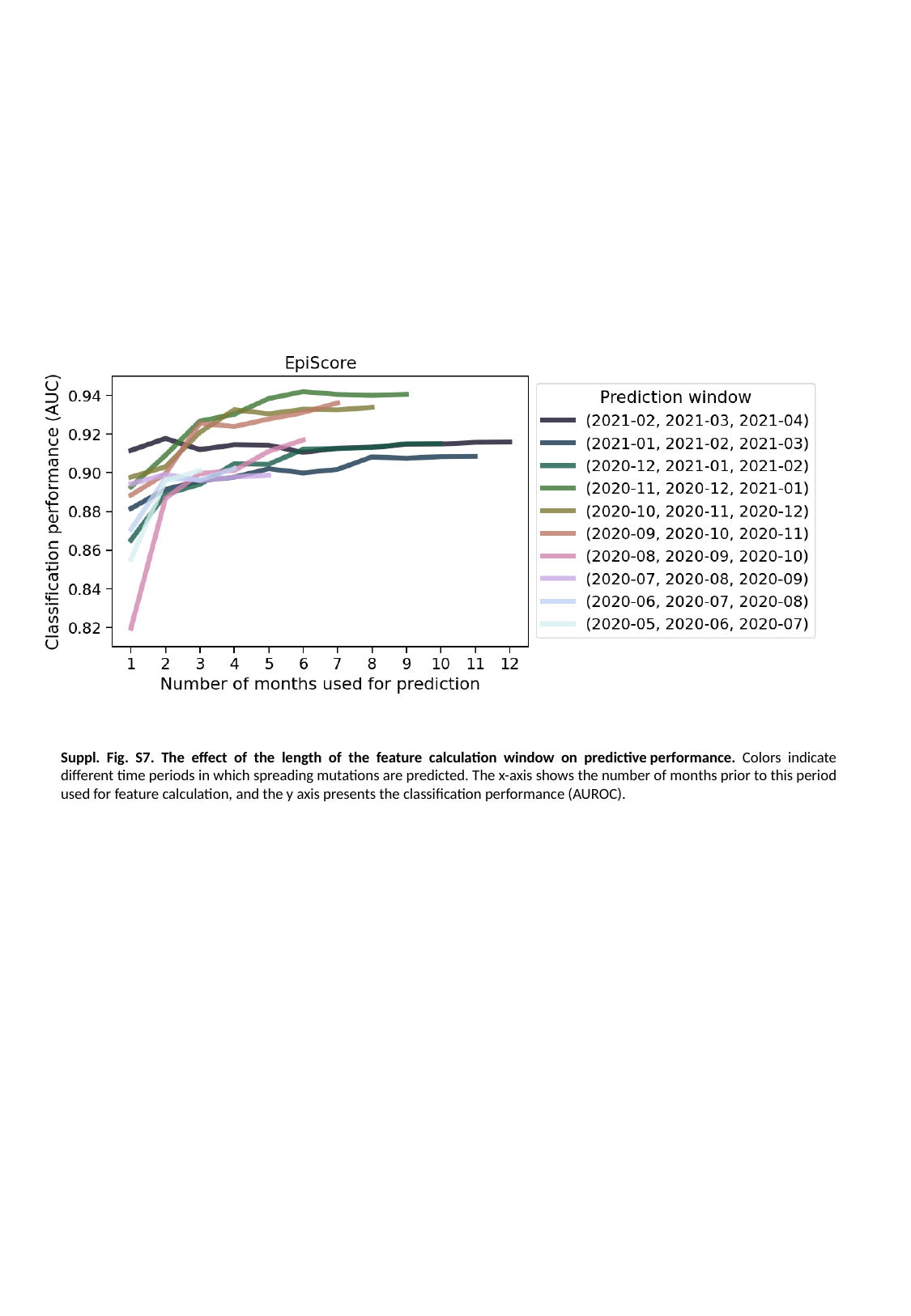

Suppl. Fig. S7. The effect of the length of the feature calculation window on predictive performance. Colors indicate different time periods in which spreading mutations are predicted. The x-axis shows the number of months prior to this period used for feature calculation, and the y axis presents the classification performance (AUROC).

### Slide 10
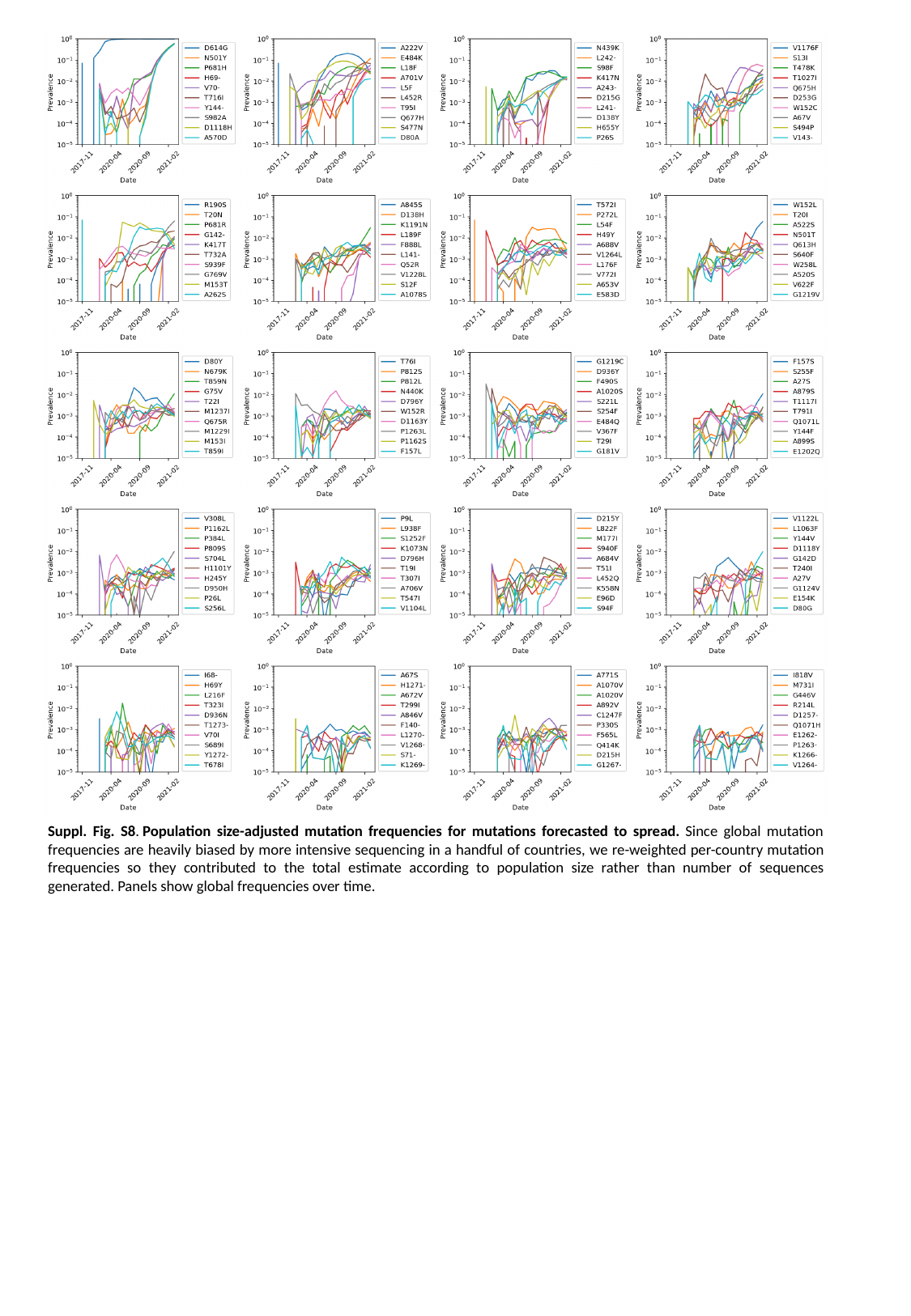

Suppl. Fig. S8. Population size-adjusted mutation frequencies for mutations forecasted to spread. Since global mutation frequencies are heavily biased by more intensive sequencing in a handful of countries, we re-weighted per-country mutation frequencies so they contributed to the total estimate according to population size rather than number of sequences generated. Panels show global frequencies over time.

### Slide 11
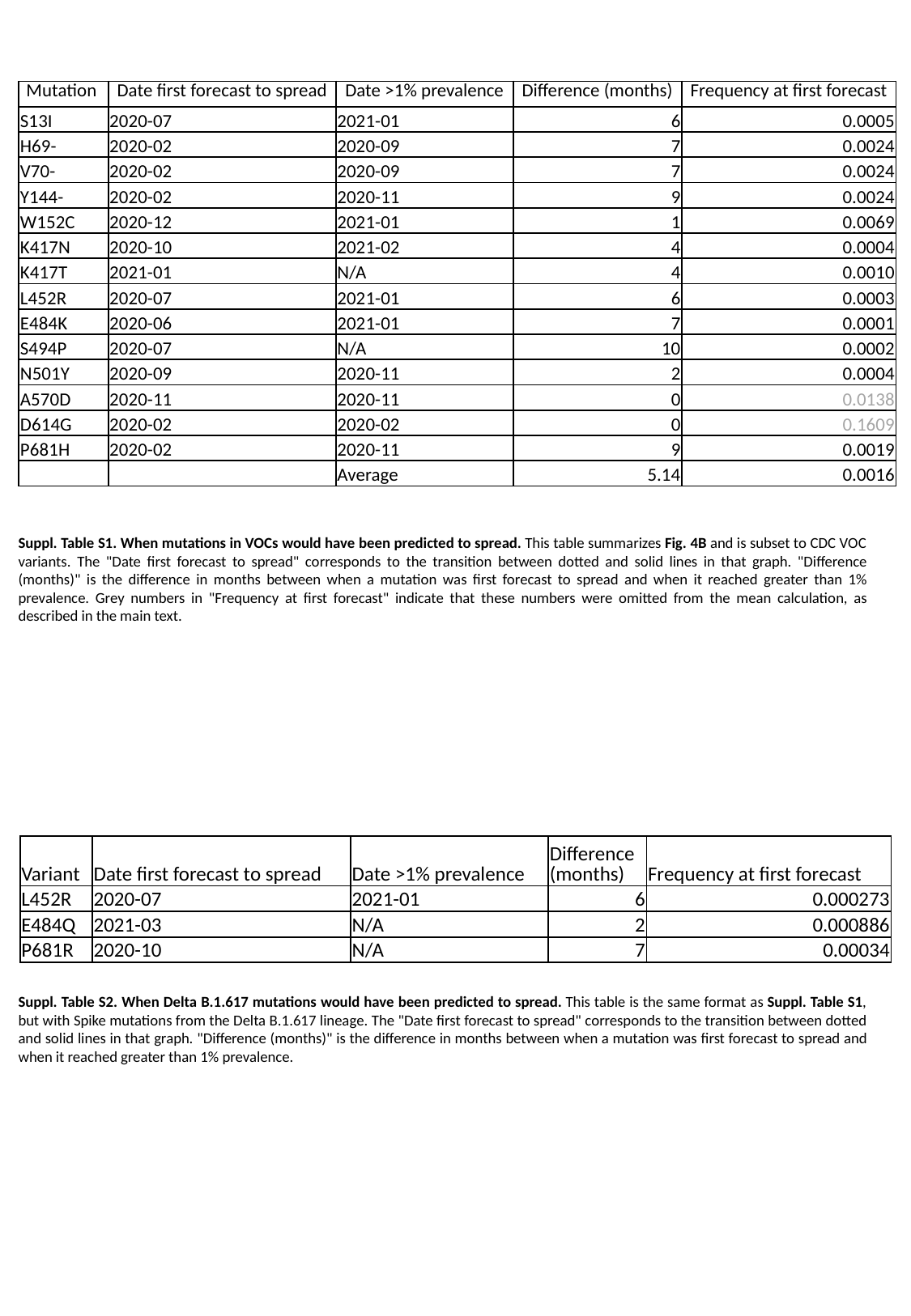

| Mutation | Date first forecast to spread | Date >1% prevalence | Difference (months) | Frequency at first forecast |
| --- | --- | --- | --- | --- |
| S13I | 2020-07 | 2021-01 | 6 | 0.0005 |
| H69- | 2020-02 | 2020-09 | 7 | 0.0024 |
| V70- | 2020-02 | 2020-09 | 7 | 0.0024 |
| Y144- | 2020-02 | 2020-11 | 9 | 0.0024 |
| W152C | 2020-12 | 2021-01 | 1 | 0.0069 |
| K417N | 2020-10 | 2021-02 | 4 | 0.0004 |
| K417T | 2021-01 | N/A | 4 | 0.0010 |
| L452R | 2020-07 | 2021-01 | 6 | 0.0003 |
| E484K | 2020-06 | 2021-01 | 7 | 0.0001 |
| S494P | 2020-07 | N/A | 10 | 0.0002 |
| N501Y | 2020-09 | 2020-11 | 2 | 0.0004 |
| A570D | 2020-11 | 2020-11 | 0 | 0.0138 |
| D614G | 2020-02 | 2020-02 | 0 | 0.1609 |
| P681H | 2020-02 | 2020-11 | 9 | 0.0019 |
| | | Average | 5.14 | 0.0016 |
Suppl. Table S1. When mutations in VOCs would have been predicted to spread. This table summarizes Fig. 4B and is subset to CDC VOC variants. The "Date first forecast to spread" corresponds to the transition between dotted and solid lines in that graph. "Difference (months)" is the difference in months between when a mutation was first forecast to spread and when it reached greater than 1% prevalence. Grey numbers in "Frequency at first forecast" indicate that these numbers were omitted from the mean calculation, as described in the main text.
| Variant | Date first forecast to spread | Date >1% prevalence | Difference (months) | Frequency at first forecast |
| --- | --- | --- | --- | --- |
| L452R | 2020-07 | 2021-01 | 6 | 0.000273 |
| E484Q | 2021-03 | N/A | 2 | 0.000886 |
| P681R | 2020-10 | N/A | 7 | 0.00034 |
Suppl. Table S2. When Delta B.1.617 mutations would have been predicted to spread. This table is the same format as Suppl. Table S1, but with Spike mutations from the Delta B.1.617 lineage. The "Date first forecast to spread" corresponds to the transition between dotted and solid lines in that graph. "Difference (months)" is the difference in months between when a mutation was first forecast to spread and when it reached greater than 1% prevalence.

### Slide 12
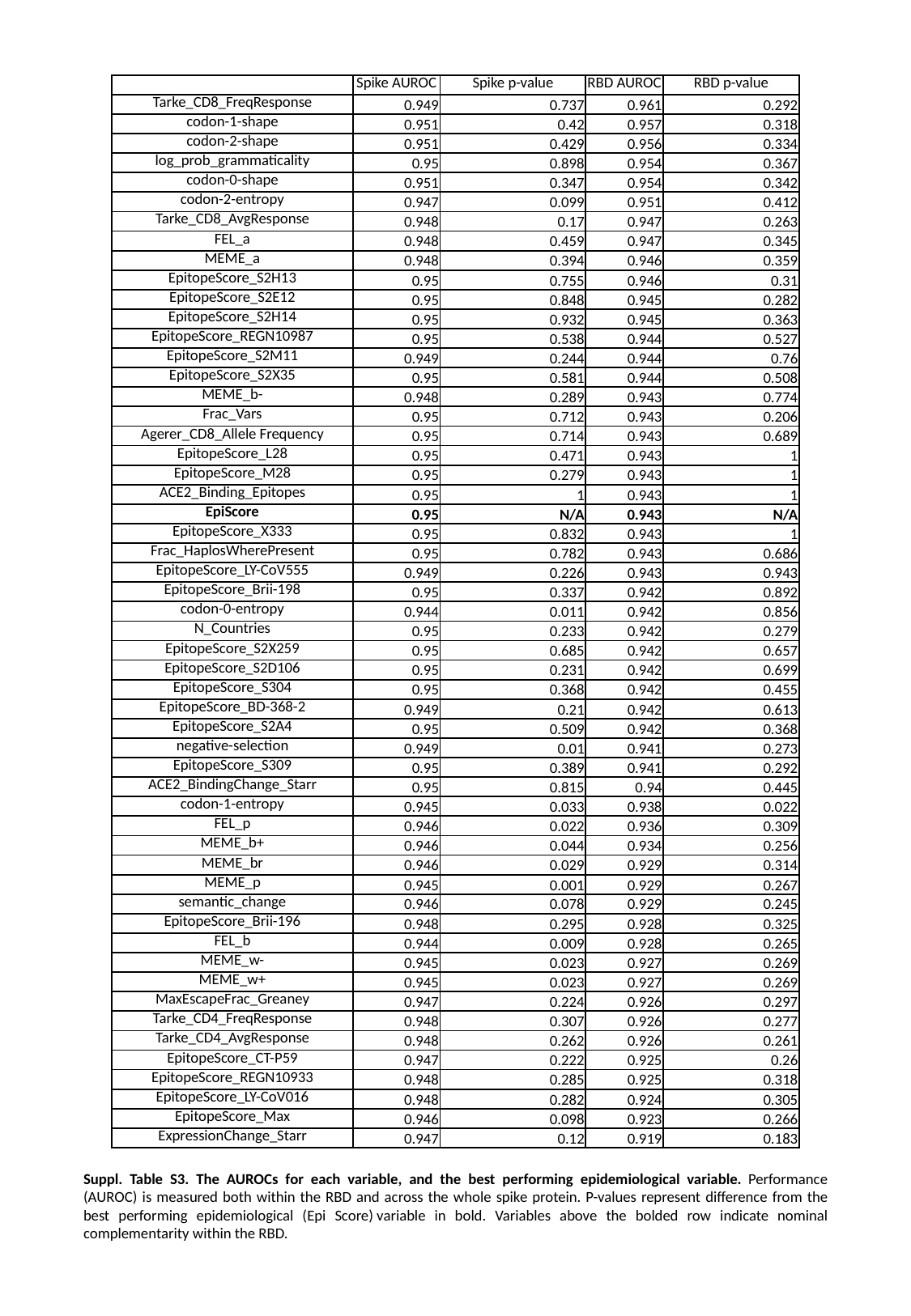

| | Spike AUROC | Spike p-value | RBD AUROC | RBD p-value |
| --- | --- | --- | --- | --- |
| Tarke\_CD8\_FreqResponse | 0.949 | 0.737 | 0.961 | 0.292 |
| codon-1-shape | 0.951 | 0.42 | 0.957 | 0.318 |
| codon-2-shape | 0.951 | 0.429 | 0.956 | 0.334 |
| log\_prob\_grammaticality | 0.95 | 0.898 | 0.954 | 0.367 |
| codon-0-shape | 0.951 | 0.347 | 0.954 | 0.342 |
| codon-2-entropy | 0.947 | 0.099 | 0.951 | 0.412 |
| Tarke\_CD8\_AvgResponse | 0.948 | 0.17 | 0.947 | 0.263 |
| FEL\_a | 0.948 | 0.459 | 0.947 | 0.345 |
| MEME\_a | 0.948 | 0.394 | 0.946 | 0.359 |
| EpitopeScore\_S2H13 | 0.95 | 0.755 | 0.946 | 0.31 |
| EpitopeScore\_S2E12 | 0.95 | 0.848 | 0.945 | 0.282 |
| EpitopeScore\_S2H14 | 0.95 | 0.932 | 0.945 | 0.363 |
| EpitopeScore\_REGN10987 | 0.95 | 0.538 | 0.944 | 0.527 |
| EpitopeScore\_S2M11 | 0.949 | 0.244 | 0.944 | 0.76 |
| EpitopeScore\_S2X35 | 0.95 | 0.581 | 0.944 | 0.508 |
| MEME\_b- | 0.948 | 0.289 | 0.943 | 0.774 |
| Frac\_Vars | 0.95 | 0.712 | 0.943 | 0.206 |
| Agerer\_CD8\_Allele Frequency | 0.95 | 0.714 | 0.943 | 0.689 |
| EpitopeScore\_L28 | 0.95 | 0.471 | 0.943 | 1 |
| EpitopeScore\_M28 | 0.95 | 0.279 | 0.943 | 1 |
| ACE2\_Binding\_Epitopes | 0.95 | 1 | 0.943 | 1 |
| EpiScore | 0.95 | N/A | 0.943 | N/A |
| EpitopeScore\_X333 | 0.95 | 0.832 | 0.943 | 1 |
| Frac\_HaplosWherePresent | 0.95 | 0.782 | 0.943 | 0.686 |
| EpitopeScore\_LY-CoV555 | 0.949 | 0.226 | 0.943 | 0.943 |
| EpitopeScore\_Brii-198 | 0.95 | 0.337 | 0.942 | 0.892 |
| codon-0-entropy | 0.944 | 0.011 | 0.942 | 0.856 |
| N\_Countries | 0.95 | 0.233 | 0.942 | 0.279 |
| EpitopeScore\_S2X259 | 0.95 | 0.685 | 0.942 | 0.657 |
| EpitopeScore\_S2D106 | 0.95 | 0.231 | 0.942 | 0.699 |
| EpitopeScore\_S304 | 0.95 | 0.368 | 0.942 | 0.455 |
| EpitopeScore\_BD-368-2 | 0.949 | 0.21 | 0.942 | 0.613 |
| EpitopeScore\_S2A4 | 0.95 | 0.509 | 0.942 | 0.368 |
| negative-selection | 0.949 | 0.01 | 0.941 | 0.273 |
| EpitopeScore\_S309 | 0.95 | 0.389 | 0.941 | 0.292 |
| ACE2\_BindingChange\_Starr | 0.95 | 0.815 | 0.94 | 0.445 |
| codon-1-entropy | 0.945 | 0.033 | 0.938 | 0.022 |
| FEL\_p | 0.946 | 0.022 | 0.936 | 0.309 |
| MEME\_b+ | 0.946 | 0.044 | 0.934 | 0.256 |
| MEME\_br | 0.946 | 0.029 | 0.929 | 0.314 |
| MEME\_p | 0.945 | 0.001 | 0.929 | 0.267 |
| semantic\_change | 0.946 | 0.078 | 0.929 | 0.245 |
| EpitopeScore\_Brii-196 | 0.948 | 0.295 | 0.928 | 0.325 |
| FEL\_b | 0.944 | 0.009 | 0.928 | 0.265 |
| MEME\_w- | 0.945 | 0.023 | 0.927 | 0.269 |
| MEME\_w+ | 0.945 | 0.023 | 0.927 | 0.269 |
| MaxEscapeFrac\_Greaney | 0.947 | 0.224 | 0.926 | 0.297 |
| Tarke\_CD4\_FreqResponse | 0.948 | 0.307 | 0.926 | 0.277 |
| Tarke\_CD4\_AvgResponse | 0.948 | 0.262 | 0.926 | 0.261 |
| EpitopeScore\_CT-P59 | 0.947 | 0.222 | 0.925 | 0.26 |
| EpitopeScore\_REGN10933 | 0.948 | 0.285 | 0.925 | 0.318 |
| EpitopeScore\_LY-CoV016 | 0.948 | 0.282 | 0.924 | 0.305 |
| EpitopeScore\_Max | 0.946 | 0.098 | 0.923 | 0.266 |
| ExpressionChange\_Starr | 0.947 | 0.12 | 0.919 | 0.183 |
Suppl. Table S3. The AUROCs for each variable, and the best performing epidemiological variable. Performance (AUROC) is measured both within the RBD and across the whole spike protein. P-values represent difference from the best performing epidemiological (Epi Score) variable in bold. Variables above the bolded row indicate nominal complementarity within the RBD.
